# NF-κB-Mediated Disruption of Key Transcription Factors Predisposes Lower Esophageal Squamous Epithelium to Esophageal Adenocarcinoma: a systematic review

**DOI:** 10.1101/2025.05.01.25326828

**Authors:** Ovais Shafi, Abdul Moez Khalid, Aakash, Moeed Ahmad, Awais Altaf Shah, Muhammad Aamir Niazi, Raveena, Faryal Yaqoob, Rahimeen Rajpar, Muhammad Hasan Aziz

## Abstract

**Objective:** The objective of this study is to determine how NF-κB-mediated transcriptional disruption of key transcription factors (TP63, SOX2, KLF4, GRHL3, ELF3, TFAP2C, RUNX1, and ZBED2) in esophageal squamous stratified epithelium leads to the Esophageal Adenocarcinoma development.

**Background:** Chronic inflammation-driven NF-κB activation disrupts key transcription factors (TP63, SOX2, KLF4, GRHL3, ELF3, TFAP2C, RUNX1, ZBED2) essential for maintaining lower esophageal squamous epithelium (SSE). This dysregulation impairs differentiation, promotes oncogenic plasticity, and enables Barrett’s metaplasia, predisposing to esophageal adenocarcinoma. Understanding these mechanisms is vital for identifying biomarkers, restoring squamous identity, and developing targeted therapies to prevent malignant transformation and improve early detection of esophageal cancer.

**Methods:** Databases, including PubMed, MEDLINE, Google Scholar, and both open-access and subscription-based journals, were searched without date restrictions to investigate how NF-κB-mediated disruption of key transcription factors (TP63, SOX2, KLF4, GRHL3, ELF3, TFAP2C, RUNX1, ZBED2) in esophageal squamous stratified epithelium predisposes it to the development of esophageal adenocarcinoma. Studies meeting the criteria outlined in the methods section were systematically reviewed to address the research question. This study adheres to the PRISMA (Preferred Reporting Items for Systematic Reviews and Meta-Analyses) guidelines.

**Results:** NF-κB disrupts key transcription factors critical for maintaining esophageal squamous identity, driving a shift toward esophageal adenocarcinoma. TP63, SOX2, and KLF4 loss weakens squamous fate, increasing plasticity. GRHL3, ELF3, and TFAP2C disruption impairs differentiation, promoting instability. RUNX1 downregulation disrupts homeostasis, while ZBED2 suppression enables columnar transformation via HNF4A activation. Chronic inflammation exacerbates these changes, fostering metaplasia and malignant progression. Understanding these mechanisms is possibly critical for developing therapies that restore transcriptional balance, block Barrett’s metaplasia, and prevent esophageal adenocarcinoma.

**Conclusion:** Esophageal adenocarcinoma arises from NF-κB-driven dysregulation of key transcription factors essential for maintaining squamous cell identity. Chronic inflammation damages TP63, SOX2, KLF4, GRHL3, ELF3, TFAP2C, RUNX1, and ZBED2, disrupting epithelial homeostasis and enabling Barrett’s metaplasia. Loss of squamous differentiation and activation of columnar pathways promote oncogenic transformation. Persistent NF-κB signaling fuels genetic instability, increasing malignancy risk. Targeting NF-κB or restoring these transcriptional regulators may prevent esophageal squamous-to-columnar transition, offering therapeutic potential against esophageal adenocarcinoma development and progression.

## Background

The transition from normal esophageal squamous epithelium (SSE) to Barrett’s esophagus and ultimately esophageal adenocarcinoma (EAC) is a critical yet poorly understood process driven by chronic inflammation. NF-κB, a central mediator of inflammation, plays a key role in altering cellular homeostasis by disrupting the balance between proliferation and differentiation in esophageal squamous progenitors [1]. The lower esophageal SSE depends on the tightly regulated activity of key transcription factors (TFs), including TP63, SOX2, KLF4, GRHL3, ELF3, TFAP2C, RUNX1, and ZBED2, to maintain stratification, lineage identity, and epithelial integrity. NF-κB-driven inflammation dysregulates these TFs, creating a permissive environment for metaplastic transformation, disrupting differentiation, and promoting oncogenic plasticity [2, 3]. TP63 and SOX2 serve as master regulators of squamous identity and proliferation, while KLF4, GRHL3, and ELF3 drive terminal differentiation and epithelial barrier function. NF-κB activation has been linked to the downregulation of these factors, shifting the balance toward an undifferentiated and proliferative phenotype susceptible to malignant transformation. RUNX1 disruption further compromises epithelial homeostasis, while ZBED2 loss enables columnar lineage conversion via HNF4A activation, a key event in Barrett’s metaplasia. TFAP2C dysregulation impairs keratinocyte differentiation, reinforcing squamous fragility [7, 8, 9]. Understanding how NF-κB-induced inflammation alters these TFs is critical for identifying early biomarkers of esophageal metaplasia and developing targeted therapies. Understanding these mechanisms could pave the way for interventions that restore squamous lineage fidelity, block malignant progression, and improve early detection of Esophageal Adenocarcinoma. This study aims to bridge critical gaps in knowledge regarding inflammation-driven transcriptional reprogramming in esophageal squamous cells, providing significant insights into carcinogenesis and potential therapeutic targets [4, 5, 6].

## Methods

### Aim of the Study

This study investigates how NF-κB-mediated disruption of key transcription factors (TP63, SOX2, KLF4, GRHL3, ELF3, TFAP2C, RUNX1, ZBED2) in esophageal squamous stratified epithelium predisposes this cell type towards Esophageal Adenocarcinoma (EAC). The primary focus is to look into:

- The NF-κB signaling in altering transcriptional regulation within esophageal squamous epithelial cells.
- How the dysregulation of specific transcription factors contributes to epithelial dedifferentiation and tumorigenic transitions.
- The link between chronic inflammation involving NF-κB activation in esophageal epithelial transformation.

### Research Question

How does NF-κB-mediated transcriptional disruption of TP63, SOX2, KLF4, GRHL3, ELF3, TFAP2C, RUNX1, and ZBED2 in esophageal squamous stratified epithelium contributes toward Esophageal Adenocarcinoma development?

### Search Focus

A comprehensive literature search was conducted using the PUBMED database, MEDLINE database, and Google Scholar, as well as open access and subscription-based journals. There were no date restrictions for published articles. The search strategy targeted:

- NF-κB-driven transcriptional regulation in epithelial cell fate transitions and its impact on the functional role of key transcription factors with focus on TP63, SOX2, KLF4, GRHL3, ELF3, TFAP2C, RUNX1, and ZBED2 in esophageal epithelium.
- NF-κB landscape underlying squamous-to-columnar transitions in the esophagus.
- NF-κB-induced epithelial reprogramming in esophageal squamous stratified epithelium towards development of esophageal adenocarcinoma.

Screening of the literature was also done on this same basis and related data was extracted. Literature search began in October 2020 and ended in October 2024. An in-depth investigation was conducted during this duration based on the parameters of the study as defined above. During revision, further literature was searched and referenced until April 2025. The literature search and all sections of the manuscript were checked multiple times during the months of revision (November 2024 – April 2025) to maintain the highest accuracy possible. This comprehensive approach ensured that the selected studies provided valuable insights with focus on the objectives of the study. This study adheres to relevant PRISMA guidelines (Preferred Reporting Items for Systematic Reviews and Meta-Analyses).

### Search Queries/Keywords

1. NF-κB and Esophageal Epithelium:

- “NF-κB signaling in esophageal epithelial transformation”
- “Chronic inflammation and NF-κB in esophageal cancer”
- “NF-κB transcriptional regulation in esophageal squamous cells”
- “Inflammatory pathways driving esophageal adenocarcinoma”
2. Key Transcription Factors in Esophageal Homeostasis & Cancer:

- “TP63 loss and esophageal adenocarcinoma”
- “SOX2 in esophageal epithelial integrity”
- “KLF4 regulation in esophageal squamous cells”
- “GRHL3 and esophageal epithelial barrier function”
- “ELF3 and esophageal cancer progression”
- “TFAP2C in esophageal epithelial differentiation”
- “RUNX1 in esophageal epithelial integrity and transformation”
- “ZBED2 and transcriptional regulation in esophageal cancer”
3. Inflammation, NF-κB, and Transcriptional Reprogramming:

- “NF-κB-driven transcriptional reprogramming in epithelial cells”
- “NF-κB in relation to each of these TFs (TP63, SOX2, KLF4, GRHL3, ELF3, TFAP2C, RUNX1, ZBED2)”
- “Chronic inflammation and metaplasia in esophagus”
- “Inflammation-induced transcriptional changes in esophageal epithelium”
- “NF-κB and loss of squamous identity in esophageal cells”

Boolean operators (AND, OR) were used to refine search results. Additional searches were conducted to identify mechanistic studies linking NF-κB activation to transcriptional disruption and epithelial reprogramming.

### Objectives of the Search

1. To determine how NF-κB activation disrupts the function of key transcription factors in esophageal squamous epithelium.
2. To investigate the role of inflammation-induced transcriptional shifts in predisposing esophageal cells to adenocarcinoma.
3. To look into the NF-κB activity with focus on esophageal epithelial dedifferentiation and tumorigenesis.

### Screening and Eligibility Criteria

Initial Screening: Titles and abstracts were reviewed to assess relevance to NF-κB signaling, transcription factor dysregulation, and EAC progression.

Full-Text Review: Studies were analyzed based on the presence of valuable insights into NF-κB-driven transcriptional reprogramming and its effects on esophageal epithelial transformation.

### Data Extraction

Key findings were categorized based on:

- NF-κB-driven changes in TP63, SOX2, KLF4, GRHL3, ELF3, TFAP2C, RUNX1, and ZBED2 expression.
- Functional consequences of transcription factor dysregulation on epithelial cell fate.
- Inflammatory signaling pathways contributing to esophageal adenocarcinoma initiation.

### Inclusion and Exclusion Criteria

Inclusion Criteria:

- Studies investigating NF-κB signaling and its role in epithelial transcription factor regulation.
- Research detailing TP63, SOX2, KLF4, GRHL3, ELF3, TFAP2C, RUNX1, and ZBED2 in esophageal squamous epithelium and cancer.
- Studies examining inflammation-induced esophageal epithelial metaplasia and carcinogenesis.
- Studies focusing on transcriptional dysregulation and epithelial reprogramming in esophageal adenocarcinoma.

Exclusion Criteria:

- Studies solely focusing on NF-κB in non-epithelial cancers.
- Research on esophageal adenocarcinoma without reference to squamous epithelial origins.
- Articles lacking insights into transcription factor dysregulation.
- Articles that did not conform to the study focus.
- Insufficient methodological rigor.

### Rationale for Screening and Inclusion

- NF-κB activation: A major driver of inflammation-induced esophageal transformation. Strongly implicated in squamous-to-columnar transitions leading to Barrett’s Esophagus and EAC.
- TP63, SOX2, KLF4, GRHL3, ELF3, TFAP2C, RUNX1, ZBED2: Essential regulators of squamous cell identity and differentiation. Transcriptional dysregulation is a key event in epithelial dedifferentiation and cancer progression.

PRISMA Flow Diagram is Fig 1.

**Fig 1.**
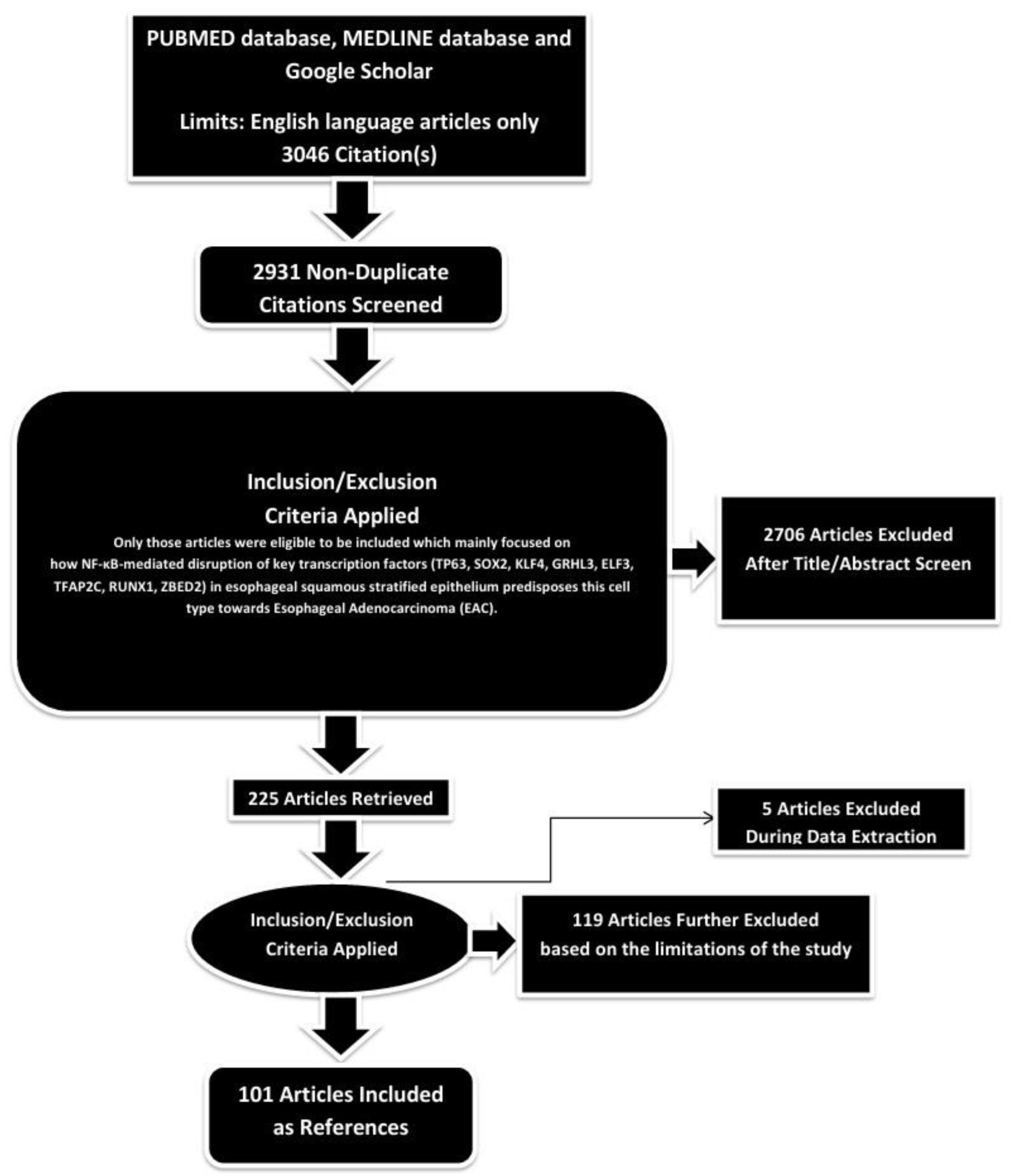
PRISMA FLOW DIAGRAM: This figure represents graphically the flow of citations in the study.

### Assessment of Article Quality and Potential Biases

Ensuring the quality and minimizing potential biases of the selected articles were crucial aspects to guarantee the rigor and reliability of the research findings.

### Quality Assessment

The initial step in quality assessment involved evaluating the methodological rigor of the selected articles. This included a thorough examination of the study design, data collection methods, and analyses conducted. The significance of the study’s findings was weighed based on the quality of the evidence presented. Articles demonstrating sound methodology— such as well-designed studies, controlled variables, and scientifically robust data—were considered of higher quality. Methodological rigor served as a significant indicator of quality.

### Potential Biases Assessment

- **Publication Bias:** To address the potential for publication bias, a comprehensive search strategy was adopted to include a balanced representation of both positive and negative results, incorporating a wide range of published articles from databases like Google Scholar.
- **Selection Bias:** Predefined and transparent inclusion criteria were applied to minimize subjectivity in the selection process. Articles were chosen based on their relevance to the study’s objectives, adhering strictly to these criteria. This approach reduced the risk of subjectivity and ensured that the selection process was objective and consistent.
- **Reporting Bias:** To mitigate reporting bias, articles were checked for inconsistencies or missing data. Multiple detailed reviews of the methodologies and results were conducted for all selected articles to identify and address any reporting bias.

By including high-quality studies and thoroughly assessing potential biases, this study aimed to provide a robust foundation for the results and conclusions presented.

### Language and Publication Restrictions

We restricted our selection to publications in the English language. There were no limitations imposed on the date of publication. Unpublished studies were not included in our analysis.

By investigating NF-κB-mediated transcriptional disruption in esophageal squamous stratified epithelium, this study provides significant insights into the mechanisms driving esophageal adenocarcinoma initiation and progression.

## Results

A total of 3046 articles were identified using database searching, and 2931 were recorded after duplicates removal. 2706 were excluded after screening of title/abstract, 119 were finally excluded, and 5 articles were excluded during data extraction.

Finally, 101 articles were included as references.

### Investigating NF-κB-mediated disruption of TFs in Esophageal Adenocarcinoma Development

#### 1. TP63

##### NF-κB-Mediated Disruption

TP63, particularly the ΔNp63α isoform, is a master regulator of stratified squamous epithelial (SSE) identity and plays a key role in the development and maintenance of the lower esophageal epithelium. During embryogenesis, TP63 is essential for basal progenitor proliferation, stratification, and the suppression of columnar differentiation programs [10]. Its expression remains critical in adulthood to preserve squamous lineage identity and prevent aberrant differentiation. However, chronic inflammation, particularly through NF-κB activation, can significantly disrupt TP63 expression, leading to pathological transitions such as Barrett’s esophagus and esophageal adenocarcinoma. NF-κB-mediated inflammation represses TP63 at multiple regulatory levels. Directly, the p65/RELA subunit of NF-κB can bind to the TP63 promoter and act as a transcriptional repressor, reducing ΔNp63α expression and compromising squamous cell maintenance. The loss of TP63 in basal progenitors increases cellular plasticity, making SSE more susceptible to metaplastic transitions. Epigenetic mechanisms further reinforce this repression. Chronic NF-κB signaling leads to DNA hypermethylation at the TP63 promoter via DNMT1 and DNMT3A, effectively silencing its transcription. Additionally, histone modifications such as EZH2-mediated H3K27me3 deposition create a repressive chromatin state, further reducing TP63 accessibility and transcription. These changes have been observed in Barrett’s esophagus and esophageal adenocarcinoma, suggesting that inflammation-induced epigenetic silencing of TP63 is a critical step in disease progression [11]. MicroRNA-mediated regulation provides another layer of TP63 suppression in response to NF-κB-driven inflammation. Inflammatory conditions upregulate miRNAs such as miR-21, miR-200, and miR-31, which directly target TP63 mRNA for degradation. miR-21, in particular, is induced by NF-κB and functions to downregulate TP63 while promoting inflammatory signaling, exacerbating the loss of squamous identity. Similarly, miR-200 family members contribute to TP63 suppression while simultaneously inducing the expression of columnar transcription factors like HNF4A and CDX2, facilitating the squamous-to-columnar transition observed in Barrett’s esophagus. The combined action of these miRNAs accelerates the loss of TP63, weakening the squamous epithelial barrier and predisposing cells to oncogenic transformation. Oxidative stress further amplifies NF-κB-driven TP63 dysregulation. Inflammatory stimuli such as bile acid reflux and chronic gastroesophageal reflux disease (GERD) generate reactive oxygen species (ROS), which stabilize NF-κB signaling, thereby prolonging TP63 suppression. Oxidative DNA damage also triggers p53-dependent TP63 degradation, reducing its protective function in epithelial renewal. Simultaneously, ROS-induced IL-6/STAT3 activation exacerbates TP63 loss while promoting a more plastic, metaplastic phenotype. The disruption of squamous homeostasis through these mechanisms provides an environment conducive to the development of Barrett’s esophagus and, ultimately, esophageal adenocarcinoma. Pro-inflammatory cytokines such as IL-1β and TNF-α contribute to the sustained repression of TP63 by maintaining NF-κB activation [12]. These cytokines promote epithelial-mesenchymal transition (EMT) and activate Wnt/β-catenin signaling, both of which further downregulate TP63 while inducing transcription factors that drive columnar differentiation. The persistent inflammatory microenvironment ensures that TP63 expression remains suppressed, leading to a progressive loss of squamous epithelial integrity. This transition from a TP63-high squamous phenotype to a TP63-low columnar-like state represents a key step in the metaplastic and neoplastic progression of esophageal disease. The clinical implications of inflammation-induced TP63 dysregulation are significant. Methylation patterns of the TP63 promoter could serve as an early biomarker for detecting Barrett’s esophagus, while elevated levels of miR-21 and miR-200 may indicate active inflammatory suppression of TP63. Therapeutic strategies targeting these pathways could potentially restore TP63 expression and preserve squamous homeostasis. Epigenetic drugs such as DNMT or EZH2 inhibitors may reactivate TP63 transcription, while NF-κB inhibitors could prevent its initial repression. Additionally, anti-inflammatory agents targeting TNF-α and IL-1β, as well as antioxidant therapies to counteract ROS-mediated TP63 degradation, may serve as protective strategies to mitigate disease progression. The restoration of TP63 function in esophageal SSE remains a promising way for preventing inflammation-driven metaplasia and carcinogenesis [13].

##### Resultant dysregulation in cell fate predisposing to Esophageal Adenocarcinoma

NF-κB-driven disruption of TP63 profoundly alters the cell fate of lower esophageal squamous cell epithelium, creating a permissive environment for Barrett’s esophagus and subsequent esophageal adenocarcinoma. TP63, particularly the ΔNp63α isoform, is essential for maintaining squamous epithelial identity by regulating basal cell proliferation, stratification, and differentiation. It acts as a key barrier against aberrant lineage transitions, ensuring that squamous cells do not adopt columnar or glandular characteristics. However, chronic inflammation, often driven by NF-κB activation in response to bile acid reflux, gastroesophageal reflux disease (GERD), and other inflammatory insults, disrupts TP63 expression at multiple regulatory levels, leading to a loss of squamous identity and increased plasticity of esophageal epithelial cells [14]. At the transcriptional level, NF-κB, particularly the p65/RELA subunit, directly binds to the TP63 promoter and represses its expression. This suppression weakens the ability of basal progenitor cells to maintain a squamous differentiation program, making them more susceptible to environmental cues that favor a columnar lineage. The loss of TP63 compromises the epithelial barrier, increasing susceptibility to further inflammatory damage and promoting a chronic wound-healing response, which further activates NF-κB. This self-reinforcing loop ensures the continuous suppression of TP63, thereby destabilizing squamous homeostasis and setting the stage for metaplastic transformation [15]. Additionally, NF-κB-induced activation of epigenetic modifiers such as DNMT1 and EZH2 results in TP63 promoter hypermethylation and H3K27me3 deposition, effectively silencing TP63 expression and reinforcing the shift away from squamous lineage commitment. MicroRNA-mediated regulation further amplifies NF-κB-driven TP63 suppression. Pro-inflammatory conditions upregulate miR-21, miR-31, and miR-200 family members, all of which directly target TP63 for post-transcriptional repression. miR-21, a well-known NF-κB-responsive miRNA, actively downregulates TP63 while promoting inflammatory and oncogenic pathways, accelerating the transition away from squamous differentiation. Simultaneously, miR-200-mediated repression of TP63 coincides with the induction of columnar transcription factors such as HNF4A and CDX2, facilitating the squamous-to-columnar transition characteristic of Barrett’s esophagus. As TP63 expression declines, basal progenitors lose their commitment to squamous fate, becoming increasingly plastic and capable of adopting a columnar identity under the influence of surrounding inflammatory and stromal signals. Oxidative stress generated by chronic inflammation further exacerbates TP63 dysregulation. Bile acid reflux and GERD-induced oxidative DNA damage contribute to NF-κB stabilization, leading to sustained suppression of TP63. Additionally, oxidative stress activates IL-6/STAT3 signaling, which cooperates with NF-κB to suppress squamous differentiation while promoting a more regenerative, metaplastic phenotype [16]. This persistent activation of inflammatory pathways drives the gradual replacement of TP63-positive squamous epithelium with columnar-like cells that express intestinal markers, a hallmark of Barrett’s esophagus. The loss of TP63 also coincides with the upregulation of pathways associated with epithelial-mesenchymal transition (EMT), further facilitating a departure from squamous differentiation and increasing the potential for neoplastic progression. As TP63 suppression continues, the esophageal epithelium becomes increasingly predisposed to esophageal adenocarcinoma. The absence of TP63-mediated cell cycle control and differentiation cues leaves basal progenitor cells vulnerable to oncogenic transformation. The emergence of Barrett’s esophagus as an intermediary step is particularly significant, as it represents a chronic inflammatory niche where columnar metaplasia persists under sustained NF-κB activity. In this altered environment, the continued influence of inflammatory cytokines such as IL-1β, TNF-α, and IL-6 maintains the repression of TP63 while simultaneously activating oncogenic drivers such as Wnt/β-catenin and Myc. This convergence of inflammatory suppression of TP63, enhanced cellular plasticity, and oncogenic activation ultimately accelerates the transition from Barrett’s esophagus to esophageal adenocarcinoma. The clinical implications of NF-κB-driven TP63 repression highlight potential therapeutic strategies aimed at restoring squamous homeostasis and preventing malignant transformation [17]. Targeting NF-κB signaling through inhibitors or anti-inflammatory agents could help maintain TP63 expression and preserve squamous identity. Epigenetic therapies that reverse TP63 silencing, such as DNMT or EZH2 inhibitors, may also restore its function and prevent metaplastic progression. Additionally, miRNA-based therapeutics that block TP63-targeting miRNAs, such as anti-miR-21 or anti-miR-200, could serve as molecular interventions to sustain squamous differentiation. Antioxidant treatments aimed at reducing oxidative stress and its impact on TP63 degradation may further mitigate the inflammatory transition toward columnar metaplasia and tumorigenesis. Collectively, understanding how NF-κB disrupts TP63 in esophageal squamous cells provides critical insights into the pathogenesis of Barrett’s esophagus and esophageal adenocarcinoma, offering a foundation for developing targeted therapeutic strategies to prevent or reverse this disease progression [18, 19].

#### 2. Sox2

##### NF-κB-Mediated Disruption

SOX2 plays a fundamental role in maintaining the squamous identity of the lower esophageal epithelium by promoting basal progenitor proliferation and restricting the activation of columnar differentiation programs. As a key transcription factor involved in stratified epithelial maintenance, SOX2 ensures that esophageal squamous cells retain their lineage identity and resist transdifferentiation into columnar or intestinal-like cells. However, inflammation, particularly chronic exposure to bile acids, gastroesophageal reflux disease (GERD), and persistent NF-κB activation, profoundly disrupts SOX2 expression, leading to cellular plasticity that predisposes the epithelium to Barrett’s metaplasia and esophageal adenocarcinoma [20]. NF-κB suppresses SOX2 transcription through direct and indirect mechanisms, altering the epigenetic landscape and favoring the loss of squamous cell identity. The NF-κB subunit p65 (RELA) can directly interact with SOX2 regulatory elements, leading to transcriptional repression through recruitment of repressive cofactors such as histone deacetylases (HDACs) and polycomb repressive complexes. Additionally, chronic inflammation induces DNA methylation at SOX2 promoter regions via upregulation of DNMT1, effectively silencing its expression in basal progenitor cells. As SOX2 levels decline, these cells lose their ability to sustain squamous differentiation, increasing their susceptibility to signals that promote intestinal metaplasia. Furthermore, NF-κB-driven activation of EZH2 facilitates H3K27me3 deposition at the SOX2 promoter, reinforcing its repression and accelerating the switch from squamous to columnar fate. MicroRNA regulation serves as another key mechanism by which inflammation suppresses SOX2 expression [21]. NF-κB-responsive miRNAs such as miR-21, miR-31, and the miR-200 family target SOX2 mRNA for degradation, further reducing its availability in esophageal epithelial cells. miR-21, a well-characterized inflammatory microRNA, is upregulated in response to chronic bile acid exposure and TNF-α signaling, leading to decreased SOX2 expression and enhanced epithelial plasticity. Simultaneously, miR-200 family members, particularly miR-200c, suppress SOX2 while promoting the expression of columnar differentiation markers such as HNF4A and CDX2. The reduction of SOX2 removes a critical barrier preventing the activation of columnar-associated transcriptional programs, shifting the esophageal epithelium toward an intestinalized state characteristic of Barrett’s esophagus. Inflammation-induced oxidative stress further exacerbates SOX2 downregulation by promoting proteasomal degradation of SOX2 protein. Bile acid reflux and GERD-associated oxidative damage enhance ROS production, leading to activation of stress-response pathways such as JNK and p38 MAPK [22]. These pathways facilitate the phosphorylation and subsequent ubiquitination of SOX2, marking it for proteasomal degradation. As SOX2 protein levels decline, basal progenitor cells become increasingly permissive to reprogramming cues from the inflammatory microenvironment, further accelerating the transition from squamous to columnar epithelium. NF-κB-mediated cytokine signaling also plays a key role in SOX2 suppression and lineage destabilization [23]. IL-6/STAT3 signaling, activated in response to chronic inflammation, synergizes with NF-κB to inhibit SOX2 expression while promoting the expression of intestinal transcription factors. IL-1β and TNF-α, both key NF-κB-driven inflammatory mediators, further contribute to this repression by activating pathways that favor wound healing and metaplastic transformation over squamous differentiation. The loss of SOX2 creates a permissive environment in which esophageal basal cells become highly plastic, allowing them to adopt columnar characteristics under sustained inflammatory pressure [24]. As SOX2 expression diminishes, esophageal epithelial cells become increasingly predisposed to Barrett’s esophagus, a well-recognized precursor to esophageal adenocarcinoma. Without SOX2, squamous progenitors lose their lineage commitment, permitting the activation of HNF4A, CDX2, and other transcriptional regulators that drive intestinal metaplasia. This transition is further reinforced by NF-κB-mediated upregulation of BMP4 and Wnt/β-catenin signaling, which establish a pro-metaplastic niche that sustains Barrett’s epithelium. The persistence of this inflammatory milieu ensures continued repression of SOX2, locking the epithelium into a columnar fate and increasing its susceptibility to oncogenic transformation. Additionally, the loss of SOX2 removes a critical tumor-suppressive mechanism, allowing aberrant proliferation and reduced cell cycle control, further accelerating neoplastic progression. Therapeutic strategies targeting NF-κB-driven SOX2 suppression could provide ways to restore squamous identity and prevent metaplastic progression. Anti-inflammatory agents that inhibit NF-κB signaling may preserve SOX2 expression and maintain squamous homeostasis. Epigenetic therapies that reverse SOX2 silencing, such as DNMT and EZH2 inhibitors, could restore its function and prevent lineage destabilization [25]. Additionally, miRNA-based therapeutics that inhibit SOX2-targeting miRNAs, such as anti-miR-21 or anti-miR-200c, may serve as potential interventions to sustain SOX2-mediated squamous differentiation. Antioxidant therapies aimed at reducing ROS-mediated degradation of SOX2 protein could further mitigate inflammatory-mediated lineage reprogramming. Ultimately, the NF-κB-driven suppression of SOX2 represents a critical early step in the transition from normal squamous epithelium to Barrett’s metaplasia and esophageal adenocarcinoma, highlighting the importance of targeting this inflammatory disruption to prevent disease progression [26].

##### Resultant dysregulation in cell fate predisposing to Esophageal Adenocarcinoma

SOX2 is a key transcription factor that maintains the squamous lineage identity of the lower esophageal epithelium by regulating basal progenitor proliferation and preventing the activation of columnar differentiation programs. Its expression is essential for preserving the stratified squamous architecture of the esophagus and resisting metaplastic transformation. However, chronic inflammation, particularly mediated by NF-κB activation, disrupts SOX2 expression and destabilizes squamous cell fate, creating a permissive environment for Barrett’s metaplasia and esophageal adenocarcinoma. NF-κB directly and indirectly represses SOX2, altering transcriptional networks, epigenetic states, and cellular signaling pathways in a way that drives a transition from squamous to columnar epithelium. NF-κB signaling suppresses SOX2 transcription through multiple mechanisms. Prolonged activation of NF-κB, triggered by chronic gastroesophageal reflux disease (GERD) or persistent bile acid exposure, leads to direct repression of SOX2 by recruiting transcriptional repressors such as histone deacetylases (HDACs) and polycomb repressive complexes [27, 28]. The NF-κB subunit RELA (p65) can bind to regulatory elements of the SOX2 promoter, promoting repressive histone modifications, including H3K27 trimethylation, which leads to long-term silencing of SOX2 in basal progenitor cells. This silencing disrupts the ability of these cells to maintain squamous differentiation and renders them more responsive to signals that favor columnar transformation. Additionally, NF-κB activation upregulates DNA methyltransferases such as DNMT1, which further contributes to the epigenetic silencing of SOX2 by depositing DNA methylation marks on its promoter, effectively shutting down its expression and leading to a loss of squamous lineage commitment. Inflammatory cytokines driven by NF-κB, such as IL-6, IL-1β, and TNF-α, further contribute to SOX2 downregulation by modulating cellular signaling pathways that favor metaplastic transformation. IL-6, through STAT3 activation, represses SOX2 expression while simultaneously promoting the expression of intestinal transcription factors such as CDX2 and HNF4A, which drive columnar differentiation [29]. TNF-α and IL-1β enhance this effect by activating MAPK and JNK pathways, which further destabilize squamous cell fate by promoting inflammatory stress responses that alter chromatin accessibility and gene expression. The resulting shift in transcriptional programs pushes the lower esophageal epithelium toward a more plastic, regenerative state that is highly susceptible to transdifferentiation into Barrett’s metaplasia. At the post-transcriptional level, NF-κB-driven inflammation induces microRNAs that directly target SOX2 for degradation. Upregulation of miR-21, a well-characterized pro-inflammatory microRNA, has been shown to suppress SOX2 expression by promoting mRNA degradation. Additionally, NF-κB activation increases levels of miR-200c, which not only represses SOX2 but also enhances the expression of columnar-associated factors, reinforcing the transition toward a non-squamous identity. The loss of SOX2 due to miRNA-mediated degradation further accelerates cellular plasticity, making the epithelium more prone to metaplastic transformation [30]. NF-κB-mediated oxidative stress also plays a key role in SOX2 destabilization. Chronic exposure to bile acids and inflammatory stimuli generates reactive oxygen species (ROS), leading to oxidative damage and activation of stress-response pathways such as p38 MAPK. These pathways trigger proteasomal degradation of SOX2, further diminishing its ability to maintain squamous differentiation. As SOX2 protein levels decline, basal progenitor cells lose the ability to resist reprogramming signals from the inflammatory microenvironment, making them increasingly susceptible to columnar differentiation cues driven by BMP4, Wnt/β-catenin, and Notch signaling. The loss of SOX2 expression is a critical early event in the development of Barrett’s esophagus, which serves as a precursor lesion for esophageal adenocarcinoma [31]. Without SOX2, the squamous epithelium loses its ability to maintain homeostasis, and basal progenitor cells become highly plastic, enabling their transdifferentiation into a columnar phenotype under the influence of NF-κB-driven inflammatory signaling. This transition is reinforced by the upregulation of BMP4 and activation of Wnt/β-catenin pathways, which establish a pro-metaplastic niche that supports the expansion of Barrett’s epithelium. Over time, this metaplastic epithelium acquires additional genetic and epigenetic alterations, including TP53 mutations and chromosomal instability, which drive its progression toward dysplasia and esophageal adenocarcinoma. The continued activation of NF-κB in this microenvironment sustains inflammatory stress, promoting further genomic instability and resistance to apoptosis, ultimately facilitating malignant transformation [32]. Restoring SOX2 expression or preventing its repression by NF-κB could serve as a potential therapeutic strategy to maintain squamous homeostasis and prevent metaplastic progression. Targeting NF-κB signaling with anti-inflammatory agents may help sustain SOX2 expression and inhibit lineage destabilization. Additionally, epigenetic therapies such as DNMT and EZH2 inhibitors could reverse the silencing of SOX2 and restore its function in maintaining squamous differentiation. MicroRNA-based interventions, including anti-miR-21 or anti-miR-200c strategies, could also be explored to prevent SOX2 degradation and preserve its tumor-suppressive function. Ultimately, NF-κB-mediated suppression of SOX2 represents a critical molecular event in the disruption of esophageal squamous epithelial fate, facilitating metaplastic transformation and predisposing the epithelium to esophageal adenocarcinoma [33].

#### 3. KLF4

##### NF-κB-Mediated Disruption

KLF4 plays a critical role in the terminal differentiation of squamous epithelial cells in the lower esophagus by promoting cell cycle exit and maintaining epithelial barrier integrity. As a transcription factor, KLF4 regulates the expression of genes involved in keratinocyte differentiation, epithelial homeostasis, and resistance to oncogenic transformation. However, its expression is highly susceptible to inflammatory signaling, particularly through NF-κB activation, which disrupts its regulatory function and predisposes the esophageal epithelium to dedifferentiation and metaplastic transformation [34]. Chronic inflammation, such as that seen in gastroesophageal reflux disease (GERD) and bile acid exposure, creates a microenvironment in which NF-κB activation directly represses KLF4 transcription, destabilizing the squamous phenotype and promoting cellular plasticity. One major mechanism by which NF-κB disrupts KLF4 expression is through direct transcriptional repression. NF-κB subunits such as RELA (p65) and p50 can interact with repressor complexes that target the KLF4 promoter, leading to histone deacetylation and chromatin remodeling that suppresses its transcription. Recruitment of histone deacetylases (HDACs) and polycomb repressive complex 2 (PRC2), which deposits H3K27 trimethylation marks, leads to the epigenetic silencing of KLF4 in basal progenitor cells of the esophageal squamous epithelium. This loss of KLF4 impairs the normal differentiation program, causing cells to remain in a proliferative, undifferentiated state, increasing the likelihood of lineage conversion under metaplastic-inducing conditions [35]. NF-κB-driven cytokines such as IL-6, TNF-α, and IL-1β further reinforce KLF4 repression through activation of STAT3 and MAPK pathways. IL-6-mediated STAT3 signaling is particularly detrimental to KLF4 expression, as STAT3 competes for enhancer binding sites that normally recruit KLF4, effectively overriding its tumor-suppressive function and promoting a regenerative, hyperplastic epithelial state. TNF-α and IL-1β enhance this effect by activating the JNK and ERK pathways, which further disrupt KLF4 expression by promoting phosphorylation-dependent degradation of KLF4 protein. This leads to a rapid decline in KLF4 levels, impairing squamous differentiation and increasing susceptibility to metaplastic transformation. At the post-transcriptional level, NF-κB signaling induces the expression of microRNAs that target KLF4 mRNA for degradation. miR-21 and miR-200c, both upregulated in chronic esophageal inflammation, directly bind to the 3′ UTR of KLF4, reducing its stability and translation [36]. This post-transcriptional repression prevents KLF4 from exerting its differentiation-promoting effects, leaving basal progenitor cells in a more plastic state where they can more easily undergo transdifferentiation into a columnar phenotype. Additionally, NF-κB-induced oxidative stress, generated by chronic bile acid exposure, promotes proteasomal degradation of KLF4 through ubiquitination pathways, further diminishing its functional presence in the esophageal epithelium. The loss of KLF4 due to NF-κB-mediated repression significantly alters esophageal epithelial cell fate. Without KLF4, squamous progenitors fail to undergo proper differentiation and remain in an undifferentiated, proliferative state. This predisposes the lower esophageal epithelium to metaplastic conversion under the influence of pro-columnar signals such as BMP4, Wnt/β-catenin, and Notch signaling, which are commonly upregulated in the setting of chronic inflammation [37]. The absence of KLF4 allows for the expansion of a Barrett’s esophagus-like cellular phenotype, characterized by columnar differentiation and the emergence of goblet cells, which serve as a precursor state for esophageal adenocarcinoma. Furthermore, KLF4 loss disrupts epithelial barrier integrity, making the esophageal lining more susceptible to further inflammatory damage and increasing exposure to genotoxic stress, which accelerates malignant progression. Restoring KLF4 expression or preventing its NF-κB-mediated repression could serve as a potential therapeutic strategy to maintain squamous homeostasis and prevent esophageal adenocarcinoma. Inhibiting NF-κB signaling through anti-inflammatory agents may help sustain KLF4 expression, preserving its role in squamous differentiation and preventing lineage destabilization. Epigenetic therapies, such as DNMT and HDAC inhibitors, could also reverse the repression of KLF4 by restoring its transcriptional activity. Additionally, targeting specific microRNAs that downregulate KLF4, such as miR-21 or miR-200c, may provide possible ways for maintaining its expression in inflamed esophageal epithelium. Ultimately, NF-κB-mediated suppression of KLF4 represents a key molecular event in the disruption of esophageal squamous epithelial fate, facilitating metaplastic transformation and predisposing the epithelium to esophageal adenocarcinoma [38, 39].

##### Resultant dysregulation in cell fate predisposing to Esophageal Adenocarcinoma

KLF4 is a critical regulator of terminal differentiation in the lower esophageal squamous epithelium, ensuring that progenitor cells exit the cell cycle and adopt a mature, stratified phenotype. Its expression is essential for maintaining epithelial integrity, suppressing uncontrolled proliferation, and preventing aberrant lineage transitions. However, in the presence of chronic inflammation, particularly through persistent activation of NF-κB signaling, KLF4 expression is significantly downregulated, leading to a loss of squamous cell identity and an increased susceptibility to metaplastic transformation [40]. NF-κB, which is activated in response to inflammatory stimuli such as bile acid reflux, microbial dysbiosis, and cytokine release, directly represses KLF4 transcription by recruiting histone deacetylases (HDACs) and polycomb repressive complexes that enforce an epigenetically repressive state. This silencing of KLF4 prevents progenitor cells from undergoing proper differentiation, leaving them in a proliferative, plastic state prone to lineage conversion. Beyond transcriptional repression, NF-κB also influences KLF4 levels through inflammatory cytokine signaling, particularly via IL-6 and TNF-α, which activate the STAT3 and MAPK pathways, respectively. IL-6/STAT3 signaling competes with KLF4 for binding at key genomic loci, disrupting its function and favoring a regenerative, hyperplastic state over normal squamous differentiation. TNF-α further exacerbates this effect by promoting JNK-mediated phosphorylation and degradation of KLF4, accelerating the loss of its tumor-suppressive functions. Additionally, NF-κB-driven oxidative stress in the esophageal epithelium contributes to proteasomal degradation of KLF4, ensuring that any residual protein is rapidly eliminated. Post-transcriptional mechanisms also reinforce KLF4 suppression, as NF-κB induces microRNAs such as miR-21 and miR-200c, which directly target KLF4 mRNA for degradation, further diminishing its levels and disrupting normal squamous differentiation [41]. The downregulation of KLF4 by NF-κB signaling has profound consequences on the fate of the lower esophageal squamous epithelium. Without KLF4 to enforce differentiation, basal progenitor cells fail to mature and instead remain in a proliferative, undifferentiated state. This creates a permissive environment for transdifferentiation, particularly under conditions that favor columnar lineage specification. In the presence of chronic inflammation and acid reflux, pro-metaplastic signaling pathways such as Wnt/β-catenin, BMP4, and Notch become upregulated, driving the replacement of squamous cells with a Barrett’s esophagus-like columnar epithelium. The loss of KLF4 removes a critical barrier to this transition, allowing progenitor cells to undergo lineage reprogramming into intestinalized columnar cells, which serve as a precursor to esophageal adenocarcinoma [42]. Moreover, KLF4 suppression weakens epithelial barrier function, exposing underlying stem cell populations to further inflammatory damage, oxidative stress, and genotoxic insults that accelerate malignant progression. As the esophageal epithelium undergoes this transition, the absence of KLF4 also disrupts key tumor-suppressive mechanisms that would otherwise prevent malignant transformation. KLF4 normally inhibits proliferation by inducing CDKN1A (p21) and suppressing MYC, yet its NF-κB-mediated repression removes this checkpoint, enabling unrestricted cell cycle progression. Additionally, KLF4 directly represses oncogenic pathways such as EMT and TGF-β signaling, both of which are activated during Barrett’s metaplasia and esophageal adenocarcinoma. Without KLF4, the esophageal epithelium not only loses its squamous identity but also gains characteristics that facilitate tumor initiation and progression. The interplay between NF-κB activation, KLF4 repression, and inflammatory signaling creates a pro-carcinogenic environment in which squamous cells gradually transition to a columnar phenotype, eventually acquiring malignant potential [43]. Ultimately, NF-κB-mediated suppression of KLF4 represents a key molecular event in the disruption of esophageal squamous epithelial homeostasis. By preventing terminal differentiation and promoting cellular plasticity, NF-κB effectively primes the esophageal epithelium for metaplastic transformation and progression toward esophageal adenocarcinoma. Targeting the NF-κB-KLF4 axis could offer a therapeutic strategy to preserve squamous identity and prevent malignant progression, either through anti-inflammatory interventions, epigenetic reprogramming, or targeted restoration of KLF4 expression in esophageal progenitor cells [44, 45].

#### 4. GRHL3

##### NF-κB-Mediated Disruption

GRHL3 is a key transcription factor involved in maintaining epithelial barrier integrity, regulating terminal differentiation, and preventing aberrant proliferation in the lower esophageal squamous epithelium. It plays a fundamental role in enforcing epithelial identity by controlling the expression of desmosomal and tight junction proteins, such as claudins and occludins, which are necessary for maintaining a functional esophageal barrier [46]. Additionally, GRHL3 suppresses pro-migratory and mesenchymal pathways, thereby restricting the transition to a more plastic and invasive phenotype. However, chronic inflammation, particularly through sustained NF-κB activation, disrupts the normal expression of GRHL3, leading to significant alterations in esophageal epithelial homeostasis that predispose cells to metaplastic transformation. The NF-κB pathway downregulates GRHL3 through multiple mechanisms, primarily by repressing its transcriptional activity. Chronic inflammation in the lower esophagus, driven by acid and bile reflux, bacterial dysbiosis, and inflammatory cytokines such as TNF-α and IL-1β, activates NF-κB signaling, which in turn recruits histone deacetylases (HDACs) and polycomb repressive complexes to the GRHL3 promoter [47]. This results in chromatin remodeling that silences GRHL3 expression, impairing its ability to maintain squamous epithelial differentiation. Additionally, NF-κB directly induces transcriptional repressors such as SNAI1 (Snail) and ZEB1, which further inhibit GRHL3 expression by promoting an epithelial-to-mesenchymal-like transition. The loss of GRHL3-driven differentiation programs weakens epithelial integrity, leaving the esophageal epithelium vulnerable to inflammatory damage and increased susceptibility to lineage plasticity. Beyond transcriptional repression, NF-κB-mediated inflammation also modulates GRHL3 through post-transcriptional and post-translational mechanisms [48]. MicroRNAs induced by NF-κB, such as miR-21 and miR-155, target GRHL3 mRNA for degradation, reducing its stability and preventing efficient translation. Furthermore, prolonged exposure to inflammatory cytokines activates proteasomal degradation pathways, which selectively target GRHL3 for breakdown, further reducing its cellular levels. Oxidative stress, a hallmark of chronic inflammation, exacerbates this effect by triggering reactive oxygen species (ROS)-mediated modifications that destabilize GRHL3 protein, impairing its function as a transcriptional regulator. The suppression of GRHL3 due to NF-κB signaling has profound consequences on esophageal epithelial cell fate. Without GRHL3 to maintain epithelial barrier integrity, basal progenitor cells become exposed to inflammatory stressors, leading to increased DNA damage, uncontrolled proliferation, and a loss of differentiation cues. This environment fosters the activation of alternative lineage programs, particularly those associated with columnar differentiation. In conditions where inflammatory signaling remains chronic, pathways such as Wnt/β-catenin and BMP4 become upregulated, driving a shift toward Barrett’s esophagus-like columnar metaplasia [49]. The absence of GRHL3 allows these pro-metaplastic signals to dominate, reinforcing the transition from squamous to columnar epithelium, which is a well-recognized precursor state for esophageal adenocarcinoma. Furthermore, GRHL3 normally functions as a tumor suppressor by directly repressing oncogenic pathways such as TGF-β signaling and EMT regulators, both of which contribute to the malignant transformation of Barrett’s esophagus into esophageal adenocarcinoma. NF-κB-mediated downregulation of GRHL3 removes these suppressive constraints, allowing for unchecked cellular proliferation, enhanced migratory capacity, and increased resistance to apoptosis. This shift not only weakens epithelial cell-cell adhesion but also facilitates the acquisition of invasive properties that are hallmarks of early cancer progression. As a result, esophageal epithelial cells that have lost GRHL3 expression due to chronic NF-κB activity are more prone to acquiring genetic and epigenetic alterations that further drive neoplastic transformation [50]. NF-κB-mediated inflammation disrupts GRHL3 expression at multiple levels, leading to the destabilization of squamous epithelial homeostasis and an increased propensity for metaplastic transformation. By silencing GRHL3, NF-κB effectively removes a critical safeguard against Barrett’s esophagus and esophageal adenocarcinoma, reinforcing a microenvironment that supports malignant progression. The role of GRHL3 in maintaining esophageal epithelial integrity, therapeutic strategies aimed at restoring its expression or counteracting NF-κB-driven suppression may offer potential ways for preventing inflammation-induced esophageal malignancies [51]

##### Resultant dysregulation in cell fate predisposing to Esophageal Adenocarcinoma

NF-κB activation in response to chronic inflammation profoundly alters the fate of lower esophageal squamous epithelium by disrupting the expression and function of GRHL3, a transcription factor essential for maintaining epithelial differentiation, barrier integrity, and homeostasis. Under normal conditions, GRHL3 enforces squamous lineage commitment by regulating the expression of desmosomal proteins, tight junction components, and differentiation-associated genes. However, persistent inflammatory signaling, particularly through NF-κB, downregulates GRHL3 at multiple levels, thereby impairing its ability to maintain epithelial integrity. This disruption weakens the protective barrier of the esophageal epithelium, exposing basal progenitor cells to further inflammatory damage and creating an environment conducive to metaplastic transformation. NF-κB suppresses GRHL3 expression primarily by recruiting histone-modifying complexes that enforce transcriptional repression. Inflammatory cytokines such as TNF-α, IL-1β, and IL-6 induce NF-κB signaling, which in turn activates chromatin remodelers like HDACs and polycomb repressive complexes that silence the GRHL3 promoter. Additionally, NF-κB-driven upregulation of transcriptional repressors, including SNAI1 (Snail) and ZEB1, further inhibits GRHL3 transcription, facilitating a loss of epithelial identity [52]. The downregulation of GRHL3 impairs the differentiation of squamous progenitor cells, leading to increased plasticity and a permissive state for transdifferentiation into columnar cell types. This shift is a critical early event in the progression toward Barrett’s esophagus, a precursor lesion for esophageal adenocarcinoma. In addition to transcriptional repression, NF-κB also modulates GRHL3 post-transcriptionally through inflammatory microRNAs such as miR-21 and miR-155, which target GRHL3 mRNA for degradation. This further reduces its expression, reinforcing the failure of differentiation programs that are necessary to maintain squamous epithelial identity. NF-κB-induced oxidative stress exacerbates this effect by increasing proteasomal degradation of GRHL3 protein, thereby diminishing its functional activity within the nucleus. As GRHL3 levels decline, the epithelial barrier weakens, leading to increased permeability and exposure of basal progenitor cells to acid and bile reflux, pro-inflammatory cytokines, and other damaging stimuli. This chronic insult promotes DNA damage accumulation and accelerates the loss of squamous differentiation. The loss of GRHL3 removes a key safeguard against the activation of pro-metaplastic signaling pathways, such as Wnt/β-catenin and BMP4, which drive columnar transformation. In the absence of GRHL3, these pathways become upregulated, promoting a shift in cell fate from squamous to intestinal-like columnar epithelium [53]. This transition is a defining feature of Barrett’s esophagus and sets the stage for malignant transformation. Furthermore, GRHL3 normally functions as a tumor suppressor by repressing oncogenic pathways such as TGF-β signaling and epithelial-mesenchymal transition (EMT) regulators. NF-κB-mediated suppression of GRHL3 eliminates these protective mechanisms, allowing for unchecked proliferation, loss of adhesion, and increased invasive potential. Without GRHL3 to counteract NF-κB-driven pro-tumorigenic signaling, esophageal epithelial cells acquire increased resistance to apoptosis and undergo genetic and epigenetic alterations that accelerate neoplastic progression [54]. As a result of NF-κB-induced GRHL3 suppression, the lower esophageal squamous epithelium undergoes a stepwise transformation, beginning with the loss of barrier function, followed by metaplastic conversion to columnar epithelium, and ultimately culminating in the development of esophageal adenocarcinoma. The chronic inflammatory environment perpetuated by NF-κB not only sustains this process but also promotes immune evasion and resistance to differentiation cues, further driving the progression toward malignancy. The critical role of GRHL3 in maintaining esophageal epithelial homeostasis, therapeutic strategies aimed at restoring its expression or counteracting NF-κB-mediated repression could serve as potential interventions to prevent Barrett’s esophagus and esophageal adenocarcinoma [55, 56, 57].

#### 5. ELF3

##### NF-κB-Mediated Disruption

ELF3, an epithelial-specific ETS transcription factor, plays role in maintaining the differentiation and barrier integrity of the lower esophageal squamous epithelium. It regulates the expression of genes involved in epithelial homeostasis, including those controlling cell adhesion, differentiation, and response to environmental stressors. During lower esophageal development, ELF3 ensures proper squamous lineage commitment and prevents aberrant cellular plasticity [58]. However, chronic inflammation, particularly through persistent NF-κB activation, disrupts ELF3 expression at multiple levels, leading to a compromised epithelial barrier and increased susceptibility to metaplastic transformation. NF-κB signaling, induced by inflammatory cytokines such as TNF-α, IL-1β, and IL-6, suppresses ELF3 transcription through direct and indirect mechanisms, creating a permissive environment for cellular dedifferentiation and lineage switching. At the transcriptional level, NF-κB interacts with repressor complexes such as SNAI1 and ZEB1, which are upregulated in response to inflammation and actively repress ELF3 expression. This repression disrupts ELF3-mediated activation of squamous differentiation genes, leading to a shift away from a stratified squamous phenotype [59]. Additionally, NF-κB can recruit histone-modifying enzymes such as HDACs and EZH2 to the ELF3 promoter, leading to chromatin condensation and reduced transcriptional accessibility. This epigenetic silencing of ELF3 prevents the activation of key epithelial differentiation programs, fostering a more plastic and regenerative state in esophageal basal cells that increases their susceptibility to metaplastic transformation. Post-transcriptionally, NF-κB-induced microRNAs such as miR-21 and miR-135b target ELF3 mRNA for degradation, further reducing its expression. These inflammatory microRNAs are upregulated in response to chronic esophageal injury and contribute to the destabilization of ELF3 transcripts, leading to a loss of its regulatory functions in epithelial integrity. Additionally, oxidative stress generated by chronic inflammation enhances proteasomal degradation of ELF3 protein, diminishing its stability and nuclear localization. This degradation impairs ELF3’s ability to activate its target genes, exacerbating the loss of squamous identity and weakening the epithelial barrier. The dysregulation of ELF3 in an NF-κB-driven inflammatory environment removes a critical barrier to pro-metaplastic signaling pathways. ELF3 normally represses pro-columnar transcription factors such as HNF4A and CDX2, which drive intestinalization of esophageal epithelium [60]. With ELF3 expression suppressed, these factors become upregulated, facilitating the replacement of squamous epithelium with columnar-like cells characteristic of Barrett’s esophagus. This transition marks a key step in the sequence of events leading to esophageal adenocarcinoma, as it creates a cellular environment more susceptible to oncogenic transformation. Furthermore, the loss of ELF3 removes a key regulatory checkpoint against uncontrolled proliferation and inflammation-induced oncogenesis. ELF3 functions as a tumor suppressor by restricting hyperplastic growth and preventing the activation of oncogenic pathways such as Wnt/β-catenin and TGF-β signaling. NF-κB-mediated suppression of ELF3 eliminates these protective effects, allowing basal progenitor cells to escape normal differentiation cues and adopt a more proliferative, stem-like phenotype. This shift in cellular behavior, combined with continued inflammatory stress and exposure to bile acid and gastric reflux, creates a microenvironment conducive to the development of esophageal adenocarcinoma [61]. As ELF3 expression is progressively lost due to NF-κB-induced repression, the esophageal epithelium undergoes a stepwise transformation, beginning with impaired differentiation, followed by the activation of metaplastic and oncogenic pathways, ultimately culminating in malignancy. The essential role of ELF3 in maintaining esophageal epithelial homeostasis, restoring its expression or counteracting NF-κB-mediated suppression could serve as a potential therapeutic strategy to prevent or slow the progression of Barrett’s esophagus and esophageal adenocarcinoma [62].

##### Resultant dysregulation in cell fate predisposing to Esophageal Adenocarcinoma

ELF3 plays role in maintaining the differentiation and epithelial barrier integrity of the lower esophageal squamous epithelium by regulating genes involved in epithelial homeostasis, adhesion, and stress responses. Its function as a key transcriptional regulator ensures the maintenance of squamous lineage identity and prevents aberrant cellular plasticity. However, chronic inflammation, particularly through persistent NF-κB activation, disrupts ELF3 expression and function at multiple levels, leading to a loss of epithelial stability and an increased susceptibility to metaplastic transformation [63]. NF-κB, induced by pro-inflammatory cytokines such as TNF-α, IL-1β, and IL-6, actively suppresses ELF3 transcription through interactions with repressor complexes such as SNAI1 and ZEB1. These repressors are upregulated in response to inflammation and directly inhibit ELF3 expression, thereby impairing its ability to activate differentiation programs essential for squamous epithelial maintenance. Furthermore, NF-κB recruits chromatin-modifying enzymes such as HDACs and EZH2 to the ELF3 promoter, resulting in epigenetic silencing and reduced transcriptional accessibility. This suppression prevents ELF3 from exerting its normal regulatory functions, creating an environment in which basal progenitor cells lose their commitment to squamous differentiation and become more plastic. Post-transcriptionally, NF-κB-induced microRNAs such as miR-21 and miR-135b target ELF3 mRNA for degradation, further reducing its expression and stability. These inflammatory microRNAs, which are highly upregulated in response to chronic esophageal injury, destabilize ELF3 transcripts and prevent translation, accelerating the loss of its epithelial protective functions [64]. Additionally, oxidative stress generated by chronic inflammation enhances the proteasomal degradation of ELF3 protein, reducing its nuclear localization and impairing its ability to activate squamous differentiation genes. The resulting downregulation of ELF3 disrupts the normal differentiation trajectory of the esophageal squamous epithelium, allowing basal progenitor cells to escape normal lineage constraints. This loss of ELF3-mediated transcriptional control removes an essential barrier to pro-metaplastic pathways, facilitating a phenotypic shift toward an intestinalized columnar-like state. With ELF3 expression suppressed, key pro-columnar transcription factors such as HNF4A and CDX2 become aberrantly upregulated, driving the initiation of Barrett’s metaplasia [65]. ELF3 normally functions as a repressor of these factors, ensuring that the esophageal epithelium remains squamous. However, in an NF-κB-driven inflammatory environment, the loss of ELF3 removes this repression, allowing the expansion of columnar progenitor populations that replace the native squamous epithelium. This metaplastic transition is a key precursor event in the development of esophageal adenocarcinoma, as the newly established columnar-like cells acquire increased susceptibility to oncogenic transformation. The loss of ELF3 also compromises epithelial barrier integrity, leading to increased permeability, chronic injury, and further inflammatory signaling, thereby sustaining a feedforward loop of NF-κB activation and ELF3 suppression [66]. This persistent inflammatory signaling promotes continuous cellular dedifferentiation, unchecked proliferation, and the accumulation of oncogenic mutations, ultimately increasing the risk of malignant transformation. As ELF3 expression is progressively lost, its tumor-suppressive functions are also diminished, allowing pathways such as Wnt/β-catenin and TGF-β signaling to become dysregulated. ELF3 normally acts as a gatekeeper against hyperplastic growth by restricting aberrant proliferative signaling, but NF-κB-mediated suppression of ELF3 removes this safeguard, enabling basal progenitor cells to acquire a more stem-like, proliferative phenotype. This shift in cellular behavior, combined with persistent inflammatory stress and exposure to bile acid and gastric reflux, creates an environment conducive to neoplastic progression. The esophageal epithelium undergoes a stepwise transformation from an impaired squamous phenotype to metaplastic Barrett’s epithelium and ultimately to esophageal adenocarcinoma [67]. The essential role of ELF3 in maintaining epithelial homeostasis, strategies aimed at restoring ELF3 expression or counteracting NF-κB-mediated repression could serve as potential therapeutic approaches to prevent or delay the progression of Barrett’s esophagus and esophageal adenocarcinoma [68, 69].

#### 6. TFAP2C

##### NF-κB-Mediated Disruption

TFAP2C plays a key role in regulating epithelial cell differentiation and maintaining the homeostasis of the lower esophageal squamous epithelium. As a transcription factor involved in keratinocyte differentiation, TFAP2C ensures the structural integrity and function of squamous epithelial cells by modulating the expression of key genes involved in epithelial identity and stratification. However, chronic inflammation, particularly through the activation of NF-κB signaling, disrupts TFAP2C gene expression at multiple levels, leading to impaired differentiation and increased susceptibility to metaplastic transformation. Inflammatory cytokines such as TNF-α, IL-1β, and IL-6, which are elevated in chronic esophageal inflammation, induce NF-κB activity, which in turn represses TFAP2C transcription through multiple mechanisms [70]. NF-κB directly interacts with repressive transcriptional complexes, including SNAI1 and ZEB1, which bind to the TFAP2C promoter and inhibit its transcription. Additionally, NF-κB recruits histone deacetylases (HDACs) and polycomb group proteins such as EZH2 to the TFAP2C locus, resulting in chromatin remodeling that silences gene expression. This epigenetic repression reduces the accessibility of TFAP2C to the transcriptional machinery, effectively downregulating its expression in response to chronic inflammation. Beyond transcriptional repression, NF-κB signaling also influences the post-transcriptional regulation of TFAP2C through inflammatory microRNAs. NF-κB-induced microRNAs, including miR-21 and miR-155, specifically target TFAP2C mRNA, leading to transcript degradation and inhibition of translation. This further diminishes the availability of TFAP2C protein, preventing its role in squamous differentiation [71]. Additionally, chronic oxidative stress generated by inflammation enhances the proteasomal degradation of TFAP2C, further reducing its stability and nuclear localization. The loss of TFAP2C expression disrupts the normal differentiation program of esophageal squamous epithelial cells, leading to an impaired epithelial barrier, increased basal cell proliferation, and a loss of commitment to the squamous lineage. As TFAP2C expression declines, the esophageal squamous epithelium becomes increasingly susceptible to aberrant cellular plasticity. Under normal conditions, TFAP2C suppresses the expression of columnar-specific transcription factors such as HNF4A and CDX2, which are essential for intestinal differentiation. However, NF-κB-mediated repression of TFAP2C removes this inhibitory control, allowing these pro-metaplastic factors to become aberrantly upregulated. This shift in gene expression promotes the transition from a squamous to a columnar phenotype, contributing to the initiation of Barrett’s metaplasia [72]. The loss of TFAP2C further weakens epithelial cohesion and barrier function, allowing increased exposure to bile acid and gastric reflux, which exacerbate chronic inflammation and sustain NF-κB activation. This creates a vicious cycle in which inflammation continuously drives TFAP2C suppression while promoting the expansion of metaplastic progenitor populations. As the esophageal epithelium undergoes progressive dedifferentiation, the absence of TFAP2C allows oncogenic pathways such as Wnt/β-catenin and TGF-β signaling to become dysregulated, promoting hyperproliferation and the accumulation of genetic mutations. The failure to maintain squamous differentiation removes an essential layer of tumor suppression, facilitating the transition from metaplastic Barrett’s epithelium to dysplastic lesions and ultimately to esophageal adenocarcinoma [73].

##### Resultant dysregulation in cell fate predisposing to Esophageal Adenocarcinoma

NF-κB activation in the lower esophageal squamous epithelium profoundly impacts TFAP2C expression, disrupting epithelial homeostasis and driving cellular reprogramming that predisposes to esophageal adenocarcinoma. As a key regulator of keratinocyte differentiation, TFAP2C ensures the maintenance of squamous lineage identity by promoting the expression of epithelial differentiation markers and suppressing pathways associated with columnar transformation. However, chronic inflammation leads to persistent NF-κB activation, which represses TFAP2C expression through direct transcriptional inhibition, epigenetic silencing, and post-transcriptional regulation. NF-κB interacts with repressive transcriptional regulators such as SNAI1 and ZEB1, which bind to the TFAP2C promoter and inhibit its transcription [74]. Simultaneously, NF-κB recruits chromatin-modifying enzymes, including histone deacetylases (HDACs) and the polycomb repressor EZH2, which facilitate the formation of a repressive chromatin environment, preventing TFAP2C transcriptional activation. This loss of TFAP2C leads to a failure in terminal squamous differentiation, disrupting the normal barrier function and structural integrity of the esophageal epithelium. Post-transcriptional mechanisms further compound TFAP2C suppression, as NF-κB-induced inflammatory microRNAs, such as miR-21 and miR-155, directly target TFAP2C mRNA, leading to degradation and translational repression [75]. In parallel, chronic oxidative stress and inflammatory signaling enhance the proteasomal degradation of TFAP2C protein, further reducing its nuclear availability. As a result, squamous progenitor cells fail to undergo proper differentiation, instead retaining an undifferentiated, plastic state that is more susceptible to metaplastic transformation. The loss of TFAP2C removes a critical barrier against ectopic activation of intestinal transcription factors, particularly HNF4A and CDX2, which are key drivers of Barrett’s esophagus. Under normal conditions, TFAP2C represses these columnar-associated factors, maintaining squamous lineage fidelity. However, its repression by NF-κB removes this restriction, allowing aberrant expression of intestinal differentiation programs, which initiate the transition from squamous to columnar epithelium. This inflammatory-induced loss of squamous identity establishes a microenvironment permissive for Barrett’s metaplasia, a well-documented precursor to esophageal adenocarcinoma [76]. As the epithelium continues to be exposed to bile acids and gastroesophageal reflux, chronic injury and repair cycles further reinforce NF-κB activation, exacerbating the suppression of TFAP2C and accelerating metaplastic progression. Without TFAP2C, squamous progenitor cells lose their ability to properly differentiate and instead contribute to an expanding population of metaplastic cells, which are more prone to oncogenic transformation. The dysregulated epithelium becomes increasingly susceptible to mutations in key tumor suppressor genes such as TP53 and CDKN2A, further driving the progression from metaplasia to dysplasia and ultimately adenocarcinoma. The disruption of TFAP2C not only facilitates the loss of squamous fate but also enhances cellular proliferation and survival, promoting a tumorigenic environment [77]. NF-κB-driven inflammation activates oncogenic signaling pathways such as Wnt/β-catenin and TGF-β, which synergize with the loss of TFAP2C to drive uncontrolled growth and invasion. The absence of TFAP2C also weakens epithelial barrier integrity, increasing susceptibility to further inflammatory damage and reinforcing the chronic activation of NF-κB signaling. This creates a self-sustaining feedback loop in which inflammation continuously suppresses TFAP2C while promoting metaplastic and neoplastic transformation. NF-κB-mediated repression of TFAP2C disrupts esophageal squamous epithelial fate by inhibiting differentiation, promoting cellular plasticity, and enabling the ectopic activation of columnar differentiation programs. This shift in cell identity removes a critical barrier to Barrett’s metaplasia and esophageal adenocarcinoma, establishing a molecular landscape that favors malignant progression. Understanding these mechanisms provides valuable insights into potential therapeutic strategies aimed at restoring TFAP2C expression or targeting NF-κB-driven inflammatory repression to prevent the development of esophageal adenocarcinoma [<u>78</u>. <u>79</u>].

#### 7. RUNX1

##### NF-κB-Mediated Disruption

RUNX1 expression ensures the proper balance between progenitor cell renewal and differentiation into mature squamous cells, preventing aberrant lineage transitions [80]. However, chronic inflammation, particularly through NF-κB activation, disrupts RUNX1 expression through multiple mechanisms, leading to epithelial instability and increased susceptibility to metaplastic transformation. NF-κB directly interferes with RUNX1 transcription by recruiting repressive cofactors such as histone deacetylases (HDACs) and polycomb repressive complexes that modify chromatin to a transcriptionally inactive state. Additionally, NF-κB-induced inflammatory cytokines such as TNF-α and IL-6 activate STAT3 and AP-1, which antagonize RUNX1 transcriptional activity by competing for binding sites or forming inhibitory complexes that block its function. Epigenetic modifications further contribute to RUNX1 silencing during chronic inflammation. NF-κB-mediated recruitment of DNMT3A and EZH2 leads to hypermethylation of the RUNX1 promoter and trimethylation of histone H3 at lysine 27 (H3K27me3), creating a repressive chromatin landscape that prevents transcriptional activation [81]. At the post-transcriptional level, inflammatory microRNAs such as miR-146a and miR-155, both of which are upregulated in response to NF-κB signaling, target RUNX1 mRNA for degradation, further diminishing its expression. Additionally, oxidative stress generated by chronic inflammation promotes the ubiquitin-proteasome degradation of RUNX1 protein, further compromising its stability and function in squamous epithelial maintenance. The loss of RUNX1 disrupts the tightly controlled differentiation program of esophageal squamous progenitors, leading to an accumulation of undifferentiated, proliferative cells that are more prone to phenotypic plasticity. Without RUNX1, squamous-specific gene expression programs are downregulated, weakening the structural integrity of the epithelium and predisposing cells to alternative lineage fates. One critical consequence is the derepression of columnar differentiation factors such as HNF4A and CDX2, which are normally inhibited by RUNX1. This shift in transcriptional control facilitates the conversion of squamous cells into a Barrett’s-like columnar phenotype, particularly in the context of chronic exposure to bile acids and gastric reflux. The inflammatory suppression of RUNX1 also enhances oncogenic signaling pathways that contribute to the progression of Barrett’s esophagus to esophageal adenocarcinoma [82]. The loss of RUNX1 removes a key regulatory checkpoint that restrains Wnt/β-catenin and TGF-β signaling, leading to uncontrolled proliferation and epithelial-mesenchymal transition (EMT). NF-κB further amplifies this process by promoting the expression of SNAI1 and ZEB1, which not only drive EMT but also reinforce RUNX1 repression through feedback inhibition. As RUNX1 levels decline, the esophageal epithelium loses its ability to maintain squamous lineage fidelity, and the unchecked activation of oncogenic pathways facilitates tumor initiation and progression. Ultimately, the NF-κB-mediated dysregulation of RUNX1 establishes a microenvironment that favors metaplastic transformation and neoplastic progression. By suppressing RUNX1, inflammation disrupts the balance between squamous differentiation and cellular plasticity, creating a permissive state for Barrett’s esophagus and esophageal adenocarcinoma. This highlights the critical role of RUNX1 in maintaining esophageal epithelial identity and the importance of targeting inflammatory pathways to restore its function and prevent disease progression [83, 84].

##### Resultant dysregulation in cell fate predisposing to Esophageal Adenocarcinoma

RUNX1 is essential for maintaining the homeostasis of the lower esophageal squamous epithelium by regulating the balance between progenitor cell renewal and differentiation. It acts as a critical transcriptional regulator that reinforces squamous lineage identity, ensuring proper epithelial stratification and function. However, chronic inflammation, particularly through NF-κB signaling, disrupts RUNX1 expression and function, leading to alterations in cell fate that predispose the epithelium to metaplastic transformation and esophageal adenocarcinoma [85]. NF-κB activation in response to persistent inflammatory stimuli, such as gastroesophageal reflux and cytokine signaling, induces the recruitment of transcriptional repressors, including histone deacetylases (HDACs) and polycomb repressive complexes, to the RUNX1 promoter, resulting in transcriptional silencing. Additionally, NF-κB-driven upregulation of inflammatory cytokines such as TNF-α and IL-6 promotes STAT3 and AP-1 activation, which antagonize RUNX1 activity either by directly repressing its transcription or by competitively binding to enhancer regions that normally regulate its expression. Epigenetic mechanisms further contribute to RUNX1 suppression under inflammatory conditions [86]. NF-κB enhances the recruitment of DNA methyltransferases (DNMTs), leading to hypermethylation of the RUNX1 promoter, thereby preventing its transcriptional activation. Concurrently, NF-κB signaling promotes the expression of EZH2, a histone methyltransferase that catalyzes the trimethylation of histone H3 at lysine 27 (H3K27me3), a repressive chromatin modification that silences RUNX1 expression. At the post-transcriptional level, inflammatory microRNAs such as miR-146a and miR-155, both of which are upregulated by NF-κB, target RUNX1 mRNA for degradation, further diminishing its expression. Additionally, inflammation-induced oxidative stress accelerates RUNX1 protein degradation through the ubiquitin-proteasome pathway, thereby reducing its stability and functional availability within the cell. The loss of RUNX1 disrupts the differentiation program of esophageal squamous progenitors, leading to a shift in cellular plasticity that favors metaplastic transformation. Normally, RUNX1 acts as a gatekeeper of squamous differentiation by repressing transcription factors such as HNF4A and CDX2, which are associated with columnar cell identity. However, NF-κB-driven suppression of RUNX1 lifts this inhibition, allowing for the aberrant activation of columnar differentiation pathways that drive the progression toward Barrett’s esophagus [87]. This process is further exacerbated by inflammatory signaling, as NF-κB promotes the expression of key drivers of metaplasia, including SOX9 and FOXA2, which reinforce the transition from squamous to columnar epithelium. The resulting loss of squamous lineage fidelity weakens the esophageal epithelial barrier, making the tissue more susceptible to additional oncogenic insults. Beyond promoting metaplasia, the inflammatory suppression of RUNX1 also enhances oncogenic signaling pathways that drive the progression from Barrett’s esophagus to esophageal adenocarcinoma. RUNX1 normally serves as a tumor suppressor by restraining Wnt/β-catenin and TGF-β signaling, both of which are implicated in uncontrolled cell proliferation and epithelial-mesenchymal transition (EMT). However, NF-κB-mediated downregulation of RUNX1 removes this protective barrier, allowing for unchecked activation of these pathways, leading to increased cellular proliferation, invasion, and loss of epithelial integrity [88]. NF-κB further reinforces this process by upregulating SNAI1 and ZEB1, which not only drive EMT but also establish feedback loops that maintain RUNX1 suppression. As a result, cells lose their squamous identity, gain mesenchymal-like properties, and acquire a highly invasive phenotype that accelerates tumor progression. Ultimately, NF-κB-mediated dysregulation of RUNX1 creates a permissive environment for esophageal squamous cells to undergo lineage infidelity and transition into a metaplastic and ultimately neoplastic state. By silencing RUNX1, inflammation disrupts the homeostatic mechanisms that maintain squamous epithelial identity, allowing for the unchecked expansion of columnar-like progenitor cells that drive Barrett’s esophagus and esophageal adenocarcinoma. This highlights the critical role of RUNX1 in preserving esophageal epithelial fate and the importance of targeting inflammatory pathways to restore its function and prevent disease progression [89, 90].

#### 8. ZBED2

##### NF-κB-Mediated Disruption

ZBED2 plays a critical role in maintaining squamous lineage identity in the lower esophageal epithelium by acting as a repressor of columnar-associated transcription factors, particularly HNF4A, which is essential for intestinal and gastric epithelial differentiation. During normal development, ZBED2 ensures that esophageal basal progenitors commit to a squamous differentiation program, preventing ectopic activation of columnar differentiation pathways that could lead to metaplastic transformation. However, chronic inflammation, particularly through NF-κB signaling, disrupts the regulation of ZBED2, leading to the loss of its suppressive function and facilitating cellular plasticity that predisposes the epithelium to Barrett’s metaplasia and esophageal adenocarcinoma. NF-κB-driven inflammatory signaling can downregulate ZBED2 through multiple mechanisms [91]. At the transcriptional level, NF-κB activation promotes the recruitment of repressive histone-modifying complexes such as EZH2, which catalyzes H3K27 trimethylation (H3K27me3) at the ZBED2 promoter, thereby silencing its expression. Additionally, NF-κB enhances the expression of inflammatory cytokines such as TNF-α and IL-1β, which activate signaling cascades that promote DNA methyltransferase (DNMT)-mediated hypermethylation of the ZBED2 promoter, further restricting its transcription. In parallel, inflammatory transcription factors such as STAT3 and AP-1 can compete for enhancer binding sites that normally facilitate ZBED2 expression, thereby repressing its transcription and allowing columnar differentiation pathways to become active. Post-transcriptionally, NF-κB-regulated microRNAs, particularly miR-21 and miR-155, can directly target ZBED2 mRNA for degradation, reducing its stability and translation [92]. These microRNAs, commonly upregulated in inflamed esophageal tissues, contribute to the loss of ZBED2 protein, further weakening its inhibitory effect on columnar-associated genes. In addition to microRNA-mediated repression, NF-κB signaling enhances proteasomal degradation of ZBED2 by promoting the upregulation of E3 ubiquitin ligases that tag ZBED2 for degradation, leading to a rapid reduction in its cellular levels. This degradation mechanism is particularly relevant under conditions of chronic reflux-induced oxidative stress, which NF-κB amplifies through the induction of reactive oxygen species (ROS)-modulating enzymes. The loss of ZBED2 due to NF-κB activity has significant consequences for esophageal epithelial cell fate [93]. Normally, ZBED2 directly represses HNF4A, preventing the activation of intestinal differentiation programs that characterize Barrett’s esophagus. However, NF-κB-mediated suppression of ZBED2 removes this inhibition, allowing HNF4A to drive the expression of downstream columnar-specific genes, including CDX2 and SOX9, which promote intestinal metaplasia. Additionally, the loss of ZBED2 leads to the derepression of Wnt/β-catenin signaling, a pathway important for stem cell maintenance and oncogenic transformation. Inflammatory cytokines such as IL-6, which are upregulated by NF-κB, further potentiate this effect by activating STAT3, which cooperates with Wnt signaling to reinforce metaplastic and neoplastic progression. Furthermore, ZBED2 loss disrupts squamous cell differentiation by altering the expression of key stratification and barrier-forming genes. Under normal conditions, ZBED2 maintains the expression of squamous-specific markers such as TP63 and GRHL3, which are essential for epithelial integrity. However, NF-κB-mediated suppression of ZBED2 results in reduced TP63 expression, impairing the proliferative capacity of basal squamous progenitor cells and facilitating a transition toward a columnar phenotype [94]. This transition is further reinforced by the inflammatory induction of TGF-β signaling, which collaborates with NF-κB to promote epithelial-mesenchymal transition (EMT) and enhance cellular plasticity, thereby increasing susceptibility to oncogenic transformation. Ultimately, NF-κB-mediated dysregulation of ZBED2 disrupts the protective mechanisms that maintain esophageal squamous identity, creating a permissive environment for metaplastic transformation and tumorigenesis. By silencing ZBED2, inflammation removes a critical repressor of columnar differentiation, enabling HNF4A-driven intestinalization of the epithelium and promoting a cellular landscape conducive to esophageal adenocarcinoma. These findings, therapeutic strategies aimed at restoring ZBED2 expression or counteracting NF-κB-driven inflammatory signaling may provide a potential way for preventing Barrett’s esophagus and its progression to malignancy [95].

##### Resultant dysregulation in cell fate predisposing to Esophageal Adenocarcinoma

NF-κB activation in the lower esophageal squamous epithelium disrupts the regulatory function of ZBED2, a transcription factor for maintaining squamous identity by repressing columnar differentiation programs. Under normal conditions, ZBED2 inhibits the expression of HNF4A, a key driver of intestinal and gastric differentiation, ensuring that squamous progenitor cells do not aberrantly activate columnar lineage pathways. However, chronic inflammation, particularly in the setting of gastroesophageal reflux disease (GERD), leads to persistent NF-κB signaling, which directly and indirectly downregulates ZBED2 expression [96]. This loss of ZBED2 removes a critical barrier against metaplastic transformation, thereby facilitating the shift from squamous to columnar cell fate and increasing susceptibility to Barrett’s esophagus and esophageal adenocarcinoma. NF-κB suppresses ZBED2 through multiple mechanisms. At the transcriptional level, NF-κB promotes the recruitment of epigenetic repressors such as EZH2, which deposits repressive H3K27me3 marks on the ZBED2 promoter, thereby silencing its expression. Additionally, NF-κB-mediated upregulation of inflammatory cytokines such as IL-6 and TNF-α triggers signaling pathways that activate DNA methyltransferases (DNMTs), leading to hypermethylation of the ZBED2 promoter and further restricting its transcription. In parallel, NF-κB interacts with inflammatory transcription factors like AP-1 and STAT3, which compete for regulatory elements that normally enhance ZBED2 expression, thereby shifting the balance toward repression. These transcriptional changes create a microenvironment where ZBED2 expression is progressively lost, allowing columnar differentiation pathways to become active. Post-transcriptionally, NF-κB promotes the degradation of ZBED2 mRNA and protein through the induction of specific microRNAs and ubiquitin-mediated proteasomal degradation [97]. NF-κB-driven upregulation of miR-21 and miR-155 targets ZBED2 mRNA for degradation, reducing its stability and translation efficiency. Furthermore, NF-κB enhances oxidative stress in esophageal epithelial cells by inducing enzymes such as NOX4, which generate reactive oxygen species (ROS). These ROS further destabilize ZBED2 protein, while NF-κB-mediated induction of E3 ubiquitin ligases accelerates its proteasomal degradation. The combined effects of transcriptional repression, microRNA-mediated silencing, and protein degradation create a sustained loss of ZBED2, facilitating a shift in cell fate. The loss of ZBED2 has profound consequences for esophageal epithelial identity. Without its repressive function, HNF4A becomes ectopically expressed, leading to the activation of CDX2 and SOX9, key transcription factors that drive intestinal differentiation. The unchecked activity of these columnar lineage factors allows basal squamous progenitor cells to adopt a more intestinal phenotype, marking the initial stages of Barrett’s metaplasia. Additionally, the loss of ZBED2 disrupts the expression of squamous-specific transcription factors such as TP63 and GRHL3, which are essential for stratification and barrier function. NF-κB-mediated inflammation further exacerbates this disruption by enhancing TGF-β signaling, which promotes epithelial-mesenchymal transition (EMT), increasing cellular plasticity and predisposing the epithelium to malignant transformation [98]. As the metaplastic epithelium continues to be exposed to inflammatory signals, NF-κB drives additional oncogenic changes that accelerate progression toward esophageal adenocarcinoma. The loss of ZBED2 removes a critical safeguard against unchecked Wnt/β-catenin signaling, a pathway that is frequently activated in Barrett’s esophagus and adenocarcinoma. Additionally, the inflammatory environment fosters genetic instability, increasing the likelihood of oncogenic mutations in tumor suppressors such as TP53, further promoting malignant transformation. The combination of NF-κB-driven ZBED2 loss, activation of columnar differentiation pathways, and chronic inflammatory stress creates a high-risk setting for the development of esophageal adenocarcinoma, highlighting the critical role of NF-κB in disrupting squamous epithelial cell fate. Understanding these mechanisms provides potential therapeutic opportunities, where targeting NF-κB signaling or restoring ZBED2 function may help prevent Barrett’s esophagus and its progression to malignancy [99, 100, 101].

## Discussion

Investigating NF-κB-mediated disruption of key transcription factors in esophageal squamous epithelium is important for understanding the mechanisms driving esophageal adenocarcinoma. TP63 serves as the master regulator of squamous epithelial fate, ensuring basal cell proliferation and stratification, but its suppression by NF-κB signaling compromises epithelial renewal, leaving cells vulnerable to fate conversion. Similarly, SOX2 maintains squamous lineage identity and actively represses Barrett’s metaplasia; its downregulation removes an essential barrier preventing the transition from squamous to columnar differentiation, a key precursor to adenocarcinoma. KLF4, which facilitates terminal differentiation of squamous cells, is also targeted by NF-κB, leading to an accumulation of undifferentiated cells with increased plasticity, a hallmark of oncogenic transformation. GRHL3, essential for epithelial barrier function and differentiation, is disrupted in inflammatory conditions, weakening cellular adhesion and promoting an environment conducive to metaplastic and dysplastic shifts. ELF3, a squamous-specific ETS transcription factor, supports epithelial differentiation, but its inhibition by NF-κB impairs the normal maturation of squamous cells, facilitating lineage instability and increased susceptibility to malignant progression. TFAP2C, which regulates keratinocyte differentiation, is similarly suppressed, leading to an erosion of squamous identity and the emergence of aberrant cell fates. RUNX1, a key regulator of esophageal epithelial homeostasis, is downregulated in response to chronic inflammation, compromising cellular turnover and disrupting normal differentiation dynamics, further exacerbating metaplastic progression. ZBED2, which inhibits HNF4A to prevent columnar transformation, is also impacted, allowing for unchecked activation of intestinal transcriptional programs that drive Barrett’s esophagus and, ultimately, esophageal adenocarcinoma. The collective disruption of these transcription factors by NF-κB not only weakens the structural integrity of the squamous epithelium but also skews cell fate determination, predisposing cells to lineage reprogramming and oncogenic transformation. The persistent inflammatory signaling associated with NF-κB exacerbates this process by promoting genetic instability and aberrant differentiation cues. Understanding these mechanistic pathways is vital for developing targeted therapies aimed at restoring squamous identity, blocking metaplastic progression, and preventing the onset of esophageal adenocarcinoma.

### Dysregulated Proliferation vs Differentiation balance in predisposing towards Esophageal Adenocarcinoma

NF-κB-induced damage in the lower esophageal squamous epithelium disrupts the delicate balance between proliferation and differentiation by altering the expression of key transcription factors that regulate epithelial homeostasis. TP63, a master regulator of basal cell proliferation and stratification, is suppressed under chronic inflammatory conditions driven by NF-κB, leading to impaired renewal of basal progenitor cells and weakening of the squamous barrier. This loss of TP63 function promotes a shift toward alternative differentiation pathways, predisposing cells to columnar metaplasia. Similarly, SOX2, which maintains squamous lineage integrity, is downregulated in response to inflammatory signaling, further compromising the proliferative capacity of squamous progenitors while permitting aberrant activation of intestinal differentiation programs characteristic of Barrett’s esophagus. The suppression of KLF4, a critical factor in promoting terminal differentiation, disrupts epithelial maturation, leaving an environment dominated by poorly differentiated, plastic cells that are more susceptible to oncogenic transformation. NF-κB signaling also interferes with differentiation pathways by targeting GRHL3, which regulates epithelial barrier function and cellular differentiation. The loss of GRHL3 weakens intercellular adhesion and structural integrity, promoting an inflammatory milieu that fosters cell fate instability. ELF3, a squamous-specific ETS factor that reinforces epithelial differentiation, is similarly repressed, impairing the expression of differentiation markers and increasing susceptibility to metaplastic conversion. The downregulation of TFAP2C, which modulates keratinocyte differentiation, further dismantles the squamous transcriptional program, allowing for a loss of squamous fate commitment. RUNX1, a key regulator of esophageal epithelial homeostasis, is also negatively affected by NF-κB, disrupting cellular turnover dynamics and weakening normal differentiation checkpoints, further accelerating metaplastic and dysplastic transitions. The inhibition of ZBED2 removes its repressive control over HNF4A, leading to activation of columnar differentiation pathways and reinforcing the transition toward Barrett’s esophagus and subsequent oncogenesis. NF-κB-driven suppression of these transcription factors skews the balance away from controlled proliferation and differentiation, favoring an environment where progenitor cells lose squamous lineage commitment while gaining plasticity toward columnar transformation. The persistence of inflammation exacerbates this process, promoting genetic instability, aberrant cell fate decisions, and an increased likelihood of esophageal adenocarcinoma development.

### Future Clinical Implications and Research Direction

Understanding how NF-κB-mediated disruption of key transcription factors contributes to the transition from esophageal squamous epithelium to adenocarcinoma has profound clinical and research implications. Clinically, this research may aid in identifying high-risk patients by using transcription factor dysregulation as biomarkers for early detection of Barrett’s esophagus and esophageal adenocarcinoma. Targeting NF-κB-driven transcriptional alterations could lead to new therapeutic strategies aimed at restoring squamous identity or preventing metaplastic progression. For instance, pharmacological inhibitors of NF-κB or agents that stabilize the expression of TP63, SOX2, and KLF4 may help maintain squamous differentiation and prevent oncogenic transformation. Additionally, the discovery of ZBED2’s role in inhibiting HNF4A suggests potential for developing therapies that reinforce squamous lineage commitment while blocking columnar reprogramming. Future research should explore how inflammation-induced transcriptional shifts interact with other oncogenic pathways, such as Wnt/β-catenin, Notch, and TGF-β signaling, to drive esophageal carcinogenesis. Investigating epigenetic modifications, such as histone methylation or DNA methylation changes in these transcription factors, may provide insights into reversible alterations that can be therapeutically targeted. Single-cell transcriptomic approaches could further elucidate how distinct epithelial subpopulations respond to chronic inflammation, identifying critical windows where intervention could prevent malignant transformation. Moreover, the development of organoid and in vivo models that replicate NF-κB-induced squamous-to-columnar transition could help evaluate targeted interventions and their ability to reverse metaplasia. As precision medicine advances, integrating transcription factor signatures into personalized risk assessments and therapeutic approaches may significantly improve outcomes for patients at risk of esophageal adenocarcinoma.

### Key Findings

By impairing differentiation programs, weakening epithelial integrity, and enhancing cellular plasticity, NF-κB creates a highly permissive environment for the development of EAC.

1. **TP63 (ΔNp63α) Suppression Weakens Squamous Identity and Promotes Columnar Transformation:** NF-κB represses TP63 (ΔNp63α), the master regulator of squamous epithelial fate, leading to the loss of basal cell proliferation control and epithelial stratification. This weakening of squamous identity creates a permissive environment for columnar metaplasia, a key precursor to EAC.
2. **SOX2 Downregulation Compromises Squamous Maintenance and Enhances Barrett’s Metaplasia Risk:** NF-κB-induced suppression of SOX2 removes a critical transcriptional safeguard against squamous-to-columnar transition. Loss of SOX2 function not only reduces squamous lineage maintenance but also enhances the likelihood of Barrett’s esophagus development, a well-established precursor to EAC.
3. **KLF4 Inhibition Shifts Proliferation-Differentiation Balance Toward a More Oncogenic State:** KLF4 is significant for terminal differentiation of squamous cells. The findings demonstrate that NF-κB represses KLF4 expression, leading to impaired differentiation, increased cellular plasticity, and heightened susceptibility to dysplastic transformation.
4. **GRHL3 Disruption Weakens Epithelial Barrier and Increases Oncogenic Potential:** GRHL3 is essential for epithelial barrier integrity and differentiation. NF-κB-mediated suppression of GRHL3 leads to compromised cell adhesion and tissue architecture, making esophageal epithelium more vulnerable to inflammatory and oncogenic signals.
5. **ELF3 Downregulation Facilitates Oncogenic Plasticity and Loss of Squamous Differentiation:** ELF3 functions as a squamous-specific ETS factor promoting epithelial differentiation. NF-κB-induced downregulation of ELF3 results in reduced squamous differentiation and increased plasticity, favoring metaplastic and dysplastic transitions.
6. **TFAP2C Dysregulation Leads to Aberrant Keratinocyte Differentiation:** TFAP2C regulates keratinocyte differentiation in squamous epithelium. NF-κB-mediated suppression of TFAP2C results in an incomplete squamous differentiation program, increasing cellular instability and susceptibility to transformation.
7. **RUNX1 Suppression Disrupts Homeostasis and Enhances Metaplastic Progression:** RUNX1 plays a key role in maintaining esophageal epithelial homeostasis. The findings indicate that NF-κB downregulates RUNX1, leading to cellular disorganization, loss of epithelial integrity, and heightened oncogenic potential.
8. **ZBED2 Repression Removes Inhibition on HNF4A, Accelerating Columnar Transformation:** NF-κB suppresses ZBED2, a key inhibitor of HNF4A. Since HNF4A is a critical driver of columnar epithelial programs, the loss of ZBED2 removes a vital barrier preventing columnar transformation, thus accelerating progression toward adenocarcinoma.

## Conclusion

The oncogenesis of esophageal adenocarcinoma (EAC) is driven by a complex interplay of inflammatory signaling, transcription factor dysregulation, and metaplastic transformation, with NF-κB playing a central role in disrupting the transcriptional landscape of the lower esophageal squamous epithelium. Under chronic inflammatory conditions, such as those induced by gastroesophageal reflux disease (GERD), persistent NF-κB activation alters the expression of key transcription factors that maintain squamous identity, including TP63, SOX2, KLF4, GRHL3, ELF3, TFAP2C, RUNX1, and ZBED2. The loss or functional impairment of these factors removes critical safeguards against aberrant cell fate decisions, facilitating the transition from a stratified squamous epithelium to a columnar metaplastic phenotype characteristic of Barrett’s esophagus, a key precursor to EAC. NF-κB-mediated suppression of TP63, a master regulator of squamous cell fate, leads to the erosion of basal cell proliferation and epithelial stratification, leaving the esophageal lining vulnerable to metaplastic conversion. Similarly, downregulation of SOX2 compromises squamous lineage maintenance and removes a critical barrier against Barrett’s metaplasia, allowing ectopic activation of intestinal differentiation pathways. The suppression of KLF4 further exacerbates this transition by impairing terminal differentiation and disrupting epithelial homeostasis, creating an environment where progenitor cells acquire increased plasticity. GRHL3, which regulates epithelial barrier integrity and differentiation, is also negatively impacted by NF-κB signaling, weakening the protective functions of the squamous epithelium and making it more susceptible to inflammatory damage and transdifferentiation. In addition to promoting metaplasia, NF-κB-driven transcriptional dysregulation accelerates oncogenic transformation by altering key epithelial differentiation programs. The suppression of ELF3, a squamous-specific ETS factor, disrupts differentiation pathways that maintain epithelial identity, allowing for the emergence of a more proliferative and less differentiated cell population prone to malignant transformation. Similarly, the inhibition of TFAP2C, which regulates keratinocyte differentiation, further dismantles the squamous transcriptional program, increasing susceptibility to columnar transformation. RUNX1, a critical factor in maintaining esophageal epithelial homeostasis, is also targeted by NF-κB, leading to unchecked inflammatory signaling, loss of epithelial integrity, and enhanced cell plasticity that facilitates oncogenesis. The downregulation of ZBED2 removes a key repressor of HNF4A, enabling the activation of columnar differentiation pathways that define Barrett’s esophagus and set the stage for neoplastic progression. Together, these NF-κB-driven disruptions create a microenvironment where the esophageal epithelium loses its ability to maintain squamous homeostasis and instead acquires traits associated with intestinal differentiation, increased proliferation, and resistance to apoptosis. The persistence of inflammatory signaling exacerbates this process, fostering genetic and epigenetic instability that promotes malignant transformation. The combined effects of transcription factor dysregulation, chronic inflammation, and aberrant cell fate reprogramming create a high-risk setting for the development of EAC, highlighting NF-κB as a central driver of disease progression. Understanding these molecular mechanisms provides valuable insight into potential therapeutic interventions, where targeting NF-κB signaling or restoring the function of key transcription factors may offer strategies to prevent or delay the onset of esophageal adenocarcinoma.

## Abbreviations

EAC: Esophageal Adenocarcinoma
NF-κB: Nuclear Factor Kappa-Light-Chain-Enhancer of Activated B Cells
TFs: Transcription Factors
TP63: Tumor Protein 63
ΔNp63α: N-Terminal Truncated Isoform of p63 Alpha
SOX2: SRY-Box Transcription Factor 2
KLF4: Kruppel-Like Factor 4
GRHL3: Grainyhead-Like Transcription Factor 3
ELF3: E74-Like ETS Transcription Factor 3
TFAP2C: Transcription Factor AP-2 Gamma
RUNX1: Runt-Related Transcription Factor 1
ZBED2: Zinc Finger, BED-Type Containing 2
HNF4A: Hepatocyte Nuclear Factor 4 Alpha
SSE: Squamous Stratified Epithelium

## Declarations

### Ethics declarations

**Ethics approval and consent to participate**

Not applicable.

### Consent for publication

Not applicable.

### Data Availability statement

All data generated or analyzed during this study are included in this article.

## Acknowledgements

We are thankful to Muhammad Danial Yaqub (MDY), MBBS, for coordinating this research project among multiple co-authors.

## Authors’ Information

1. **Ovais Shafi (OS)\*** is the author of the study and was involved in the idea, concept, design, and methodology of the study, literature search and references. He did the writing, editing, and revision of the manuscript. He was involved in drawing the findings, results, conclusions, implications of the study, interpretation of the data and was involved in all aspects of the study. He prepared and wrote discussion, results, conclusions and all areas of the study. OS extracted and analyzed the data. He was involved in critical evaluation, audit of every aspect of the study, data extraction, adherence of the study to relevant PRISMA guidelines, limitations of the study, references, and all others. He was involved in drawing PRISMA Flow Diagram. The author read and approved the manuscript. He investigated the NF-κB-mediated disruption of key transcription factors in Esophageal Adenocarcinoma development for their roles in relation to this study: TP63, SOX2, KLF4, GRHL3, ELF3, TFAP2C, RUNX1, and ZBED2. **Ovais Shafi (OS)\***, MBBS - Sindh Medical College - Dow University of Health Sciences, Karachi, Pakistan. He aspires to become an eminent ‘Physician Scientist’. He is devoted to the research in disease development mechanisms, disease origins and therapeutics. OS is also passionate about multiple research areas including clinical trials, clinical medicine, therapeutics, regenerative medicine, precision medicine including gene therapies, finding disease specific targets for gene therapy, role of disease genomics and epigenetics in diagnosis, management, and therapeutics development. He is dedicated to the field of research and clinical medicine. Email address*:
2. **Abdul Moez Khalid (AMK)** is the co-author of the study. He contributed to the results and conclusions of the study, also contributed to the writing and editing of these sections. He also contributed to the references. He contributed to investigating the NF-κB-mediated disruption of key transcription factors in Esophageal Adenocarcinoma development for their roles in relation to this study: TP63, SOX2, KLF4, GRHL3, ELF3, TFAP2C, RUNX1, and ZBED2. Abdul Moez Khalid, MBBS, currently working at Lincoln County Hospital, Lincoln, United Kingdom.
3. **Aakash (AA)** is the co-author of the study. He contributed to the results and conclusions of the study, also contributed to the writing and editing of these sections. He also contributed to the references. He contributed to investigating the NF-κB-mediated disruption of key transcription factors in Esophageal Adenocarcinoma development for their roles in relation to this study: TP63, SOX2, KLF4, GRHL3, ELF3, TFAP2C, RUNX1, and ZBED2. Aakash MD, currently working in Florida State University Cape Coral Hospital and he is MBBS from Sindh Medical College - Dow University of Health Sciences, Karachi, Pakistan. He is passionate about pursuing fellowship in gastroenterology. His goal is also to translate emerging findings in research of disease development mechanisms/ origins into their clinical implications.
4. **Moeed Ahmad (MA)** is the co-author of the study. He contributed to the results and conclusions of the study, also contributed to the writing and editing of these sections. He also contributed to the references. He contributed to investigating the NF-κB-mediated disruption of key transcription factors in Esophageal Adenocarcinoma development for their roles in relation to this study: TP63, SOX2, KLF4, GRHL3, ELF3, TFAP2C, RUNX1, and ZBED2. Moeed Ahmad, MBBS, currently working at Lincoln County Hospital, Lincoln, United Kingdom.
5. **Awais Altaf Shah (AAS)** is the co-author of the study. He contributed to the results and conclusions of the study, also contributed to the writing and editing of these sections. He also contributed to the references. He contributed to investigating the NF-κB-mediated disruption of key transcription factors in Esophageal Adenocarcinoma development for their roles in relation to this study: TP63, SOX2, KLF4, GRHL3, ELF3, TFAP2C, RUNX1, and ZBED2. Awais Altaf Shah, MBBS, currently working at Lincoln County Hospital, Lincoln, United Kingdom.
6. **Muhammad Aamir Niazi (MAN)** is the co-author of the study. He contributed to the results and conclusions of the study, also contributed to the writing and editing of these sections. He also contributed to the references. He contributed to investigating the NF-κB-mediated disruption of key transcription factors in Esophageal Adenocarcinoma development for their roles in relation to this study: TP63, SOX2, KLF4, GRHL3, ELF3, TFAP2C, RUNX1, and ZBED2. Muhammad Aamir Niazi, MBBS, currently working at Lincoln County Hospital, Lincoln, United Kingdom.
7. **Raveena (RA)** is the co-author of the study. She contributed to the results and conclusions of the study, also contributed to the writing and editing of these sections. She also contributed to the references. She contributed to investigating the NF-κB-mediated disruption of key transcription factors in Esophageal Adenocarcinoma development for their roles in relation to this study: TP63, SOX2, KLF4, GRHL3, ELF3, TFAP2C, RUNX1, and ZBED2. Raveena, MBBS - Sindh Medical College – Jinnah Sindh Medical University, Karachi, Pakistan. She is passionate about research in surgery and disease development mechanisms including neurodegenerative diseases, oncogenesis and others. She is ECFMG Certified. She is passionate about residency in Internal Medicine/Surgery. Her goal is to make significant impact in the field of Research.
8. **Faryal Yaqoob (FY)** is the co-author of the study. She contributed to the results and conclusions of the study, also contributed to the writing and editing of these sections. She also contributed to the references. She contributed to investigating the NF-κB-mediated disruption of key transcription factors in Esophageal Adenocarcinoma development for their roles in relation to this study: TP63, SOX2, KLF4, GRHL3, ELF3, TFAP2C, RUNX1, and ZBED2. Faryal Yaqoob, Doctor of Pharmacy - Faculty of Pharmacy and Pharmaceutical Sciences, Ziauddin University, Karachi Pakistan. Her goal is also to translate emerging findings in research of disease development mechanisms/ origins into their therapeutic implications.
9. **Rahimeen Rajpar (RR)** is the co-author of the study. She contributed to the results and conclusions of the study, also contributed to the writing and editing of these sections. She also contributed to the references. She contributed to investigating the NF-κB-mediated disruption of key transcription factors in Esophageal Adenocarcinoma development for their roles in relation to this study: TP63, SOX2, KLF4, GRHL3, ELF3, TFAP2C, RUNX1, and ZBED2. Rahimeen Rajpar, MD is a dedicated medical professional with a passion for unraveling the mysteries of disease origins and progression. Currently pursuing her residency in internal medicine. RR is committed to advancing the field of medical research. Her goal is to become a leader in the field of Medicine, looking at the medical intricacies from a different lens. Apart from research RR remains dedicated to improving the lives of her patients through comprehensive care and by working towards scientific discoveries. RR is a MBBS graduate from Sindh Medical College – Jinnah Sindh Medical University, Karachi, Pakistan.
10. **Muhammad Hasan Aziz (MHA)** is the author of the study. He contributed to the results and conclusions of the study, also contributed to the writing and editing of these sections. He also contributed to the references. He contributed to investigating the NF-κB-mediated disruption of key transcription factors in Esophageal Adenocarcinoma development for their roles in relation to this study: TP63, SOX2, KLF4, GRHL3, ELF3, TFAP2C, RUNX1, and ZBED2. Muhammad Hasan Aziz, Doctor of Pharmacy – FUUAST, and Masters of Business Administration - Bahria University Karachi Campus. He is passionate about research in disease development mechanisms with focus on new therapeutic implications. *The work and contributions of everyone have been described in detail, the order is randomized and the numbering is just for referencing purpose*.

